# A generic method and software to estimate the transmission advantage of pathogen variants in real-time : SARS-CoV-2 as a case-study

**DOI:** 10.1101/2021.11.26.21266899

**Authors:** Sangeeta Bhatia, Jack Wardle, Rebecca K Nash, Pierre Nouvellet, Anne Cori

## Abstract

Recent months have demonstrated that emerging variants may set back the global COVID-19 response. The ability to rapidly assess the threat of new variants in real-time is critical for timely optimisation of control strategies.

We extend the EpiEstim R package, designed to estimate the time-varying reproduction number (*R*_*t*_), to estimate in real-time the effective transmission advantage of a new variant compared to a reference variant. Our method can combine information across multiple locations and over time and was validated using an extensive simulation study, designed to mimic a variety of real-time epidemic contexts.

We estimate that the SARS-CoV-2 Alpha variant is 1.46 (95% Credible Interval 1.44-1.47) and 1.29, (95% CrI 1.29-1.30) times more transmissible than the wild type, using data from England and France respectively. We further estimate that Beta and Gamma combined are 1.25 (95% CrI 1.24-1.27) times more transmissible than the wildtype (France data). All results are in line with previous estimates from literature, but could have been obtained earlier and more easily with our off-the-shelf open-source tool.

Our tool can be used as an important first step towards quantifying the threat of new variants in real-time. Given the popularity of EpiEstim, this extension will likely be used widely to monitor the co-circulation and/or emergence of multiple variants of infectious pathogens.

**Significance Statement:** Early assessment of the transmissibility of new variants of an infectious pathogen is critical for anticipating their impact and designing appropriate interventions. However, this often requires complex and bespoke analyses relying on multiple data streams, including genomic data. Here we present a novel method and software to rapidly quantify the transmission advantage of new variants. Our method is fast and requires only routinely collected disease surveillance data, making it easy to use in real-time. The ongoing high level of SARS-CoV-2 circulation in a number of countries makes the emergence of new variants highly likely. Our work offers a powerful tool to help public health bodies monitor such emerging variants and rapidly detect those with increased transmissibility.

## Introduction

The SARS-CoV-2 pandemic has highlighted the potentially dramatic influence that emerging novel pathogen variants can have on transmission dynamics and on the control measures needed to mitigate the epidemic burden. The emergence of the Alpha variant of SARS-CoV-2 in September 2020, and of the Delta variant in December 2020 drastically altered the trajectory of the COVID-19 epidemic in several countries leading to renewed imposition of public health measures such as lockdowns [1, 2]. The continued high level of transmission of SARS-CoV-2 globally makes the emergence of new variants very likely. As of December 2021, the World Health Organization has classified four variants of SARS-CoV-2 as “variants of concern” (i.e. Alpha, Beta, Gamma and Delta), because of their increased transmissibility, severity, and/or immune escape properties compared to the circulating SARS-CoV-2 variants [3].

Rapidly quantifying characteristics of such emerging variants is critical to anticipate their potential impact and adjust interventions accordingly. Shortly after the emergence of the Alpha variant in England in September 2020 [4], a number of studies aimed to estimate its transmission potential, compared to the previously circulating non-VOC lineages [4–7]. More recently, several papers have evaluated the transmissibility of VOCs compared to non-VOC lineages [8–16]. All of these studies have developed new approaches to estimate the transmission advantages of new VOCs, often synthesising evidence from multiple data sources including genomic data. The time and expertise required to design and implement such approaches, with methods tailored to the specificity of each dataset and context, greatly limit their widescale and real-time use.

In this study, we present a new Bayesian inference method, MV-EpiEstim (for Multi-Variant EpiEstim), to estimate in real-time the transmission advantage of a new variant of a pathogen compared to a reference variant, using simple data consisting of the time series of incidence of cases of each variant in one or more locations. In the rest of the manuscript, we refer to different “variants” but the method can be equally applied to different strains. We present the method for one reference and one new variant, but the method naturally extends to more than one new variant. Our work build on a previously published methodology [17, 18] to estimate the instantaneous reproduction number *R*_*t*_ (defined as the average number of secondary cases that an individual infected at time *t* would generate if conditions remained the same as at time *t*).

We assume that locally, the transmissibility of all variants follows the same temporal pattern, i.e. the reproduction number of the new variant is the same as that of the reference variant, albeit with a multiplicative factor. We refer to this multiplicative factor as the “effective transmission advantage” of the new variant, compared to the reference variant. We further assume that the effective transmission advantage remains constant over time and across all locations under consideration.

We provide an open source implementation of our method in the R package EpiEstim [19]. The approach, which we validate on an extensive simulation study, is computationally efficient as it takes advantage of an analytical formulation of marginal posterior densities of the instantaneous reproduction number for the reference variant on the one hand, and the transmission advantage of the new variant on the other hand.

We retrospectively estimate the effective transmission advantage of the Alpha SARS-CoV-2 variant compared to the other non-VOC lineages circulating at the time using data from England and France. We show that the estimates from our method are consistent with those from several bespoke studies, and could have been obtained earlier and with improved accuracy. Our inference framework and open source software should allow rapid quantification of the effective transmission advantage of future new variants in real-time.

## Results

### Transmission advantage of SARS-CoV-2 variants

We used MV-EpiEstim to retrospectively estimate the transmission advantage of SARS-CoV-2 variants using data from England and France. The Alpha variant originated in late summer to early Autumn 2020 in England (before vaccination was initiated), where it became dominant in early 2021 (Fig. 1A). England never experienced substantial transmission of the Beta and Gamma variants, first detected in South Africa and Brazil respectively [3].

**Figure 1:**
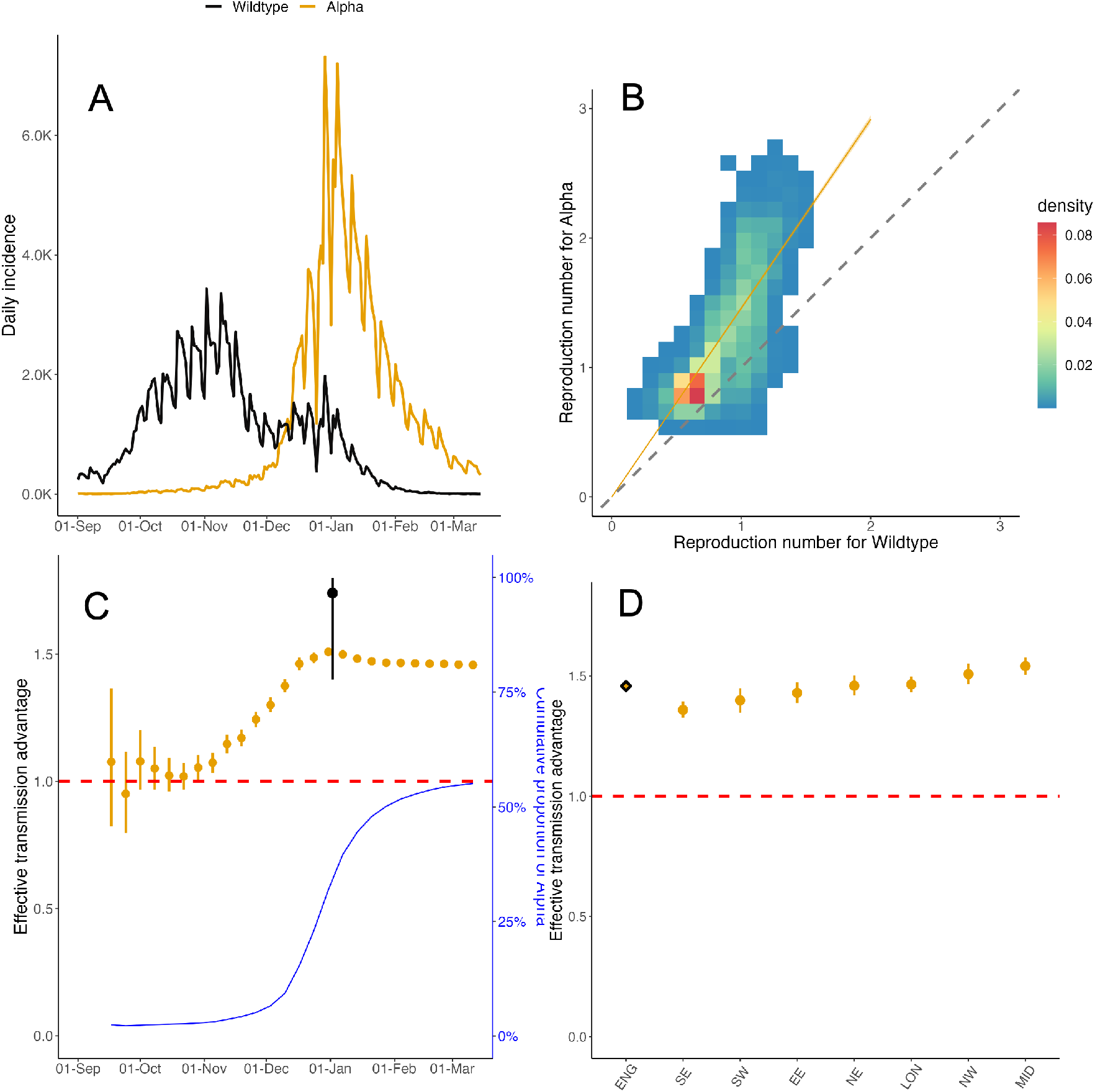
Effective transmission advantage of the Alpha SARS-CoV-2 variant over the wild-type in England. (A) The daily reported incidence of cases of the wildtype (black) and Alpha (orange) in England from September 2020 to March 2021. (B) The effective reproduction number *R*_*t*_ estimated independently for the wildtype (x-axis) and Alpha (y-axis) on sliding weekly windows. The colour of the cells indicates the density of the draws from the respective posterior distributions of *R*_*t*_. The dashed diagonal line indicates the *x* = *y* threshold. Coloured cells lying above the diagonal line suggest that Alpha is more transmissible. The orange line denotes the median effective transmission advantage estimated using MV-EpiEstim. 95% CrI were so narrow that they could not be distinguished from the line. (C) Effective transmission advantage estimated using MV-EpiEstim using data available up to the date specified on the x-axis. The dark blue line denotes the proportion of cumulative incidence of Alpha (right y-axis) counted from 1^st^ September 2020. The black estimate corresponds to the multiplicative transmission advantage of Alpha estimated by Volz et al [33] in a report published on 31^st^ December 2020. (D) Effective transmission advantage estimated using MV-EpiEstim for all NHS England regions together (diamond) and separately (solid circles), using data from 1^st^ September 2020 to 14^th^ March 2021. The NHS England regions are - East of England (EE), London (LON), Midlands (MID), North-East (NE), North-West (NW), South-East (SE), South-West (SW). In panels (C) and (D), the solid circles denote the median estimate, the vertical lines indicate the 95% CrI, and the red dashed line denotes the *ϵ* = 1 threshold.

In France, the Alpha variant emerged in early 2021, rapidly dominating cases in metropolitan France and the French West Indies [20, 21]. The Beta and Gamma variants were also circulating from January 2021 in most regions, and accounted for the majority of cases in French Guyana and la Réunion from spring 2021 [22] (Fig. S2).

We considered daily variant-specific incidence data from 7 National Health Service (NHS) regions in England between 1^st^ September 2020 and 14^th^ March 2021 (Fig. S1), and from 18 ADM2 regions in France between 18^th^ February and 30^th^ May 2021 (Figs. S2 and S3).

For simplicity, we refer to all lineages of SARS-CoV-2 other than the VOCs that were circulating at the time as ‘wildtype’. *R*_*t*_ estimates obtained independently for the wildtype and for Alpha indicated that Alpha was more transmissible (Fig. 1B). However, the magnitude of the transmission advantage (naively estimated as the ratio between the two *R*_*t*_s, see Suppl Sec. 3 for details) varied over time and across regions. Pooling these naïve estimates over time and regions yielded a highly uncertain and non-significant transmission advantage of 1.41 (95% Credible Interval (CrI) 0.86-2.01) for Alpha compared to the wildtype in England.

By explicitly assuming that the effective transmission advantage, which we denote as *ϵ*, remains constant over time and across regions, MV-EpiEstim reduces the uncertainty in the estimates. Using MV-EpiEstim with data from all NHS regions, we found strong evidence that Alpha was more transmissible than the wildtype (*ϵ* = 1.46, 95% CrI 1.44-1.47. See also Suppl Tab. S1).

To mimic real-time use, we examined how *ϵ* estimates varied as more data became available. The central estimate steadily increased from around 1 to approximately 1.5 by early December 2020, with uncertainty decreasing in that period; estimates then remained relatively stable (Fig. 1C). As a comparison, Volz et al. first estimated the multiplicative transmission advantage of Alpha to be 1.74 (95% CrI 1.40-1.80), in a report dated December 31^st^ 2020.

Alpha cases accounted for less than 10% of all cases between mid-September and early-December (Fig. 1C). The variability in *ϵ* estimates in this period suggests that accurate estimation of the transmission advantage can only be achieved once enough cases of the new variant have been observed.

We also used MV-EpiEstim to estimate *ϵ* separately for each NHS region, highlighting minor regional differences with *ϵ* ranging from 1.36 (95% CrI 1.33-1.39) in the South-East to 1.54 (95% CrI 1.50-1.58) in the Midlands (Fig. 1D, Suppl Tab. S1).

We estimated a similar, albeit slightly lower, effective transmission advantage for Alpha using data from the 18 ADM2 regions in France (*ϵ* = 1.29, 95% CrI 1.29-1.30, see Fig. S3). In region-specific analyses (excluding regions where Beta/Gamma were dominant), *ϵ* varied from 1.21 (95% CrI 1.20-1.23) in Île-de-France to 1.41 (95% CrI 1.37-1.46) in Bourgogne-Franche-Comté (Fig. S3 and Suppl Tab. S2).

Following the same approach, and using data from France, we demonstrated that the Beta and Gamma variants (combined) are also more transmissible than the wildtype (*ϵ* = 1.25, 95% CrI 1.24-1.27, Fig. S4 and Suppl Tab. S3).

### Method validation

We assessed the validity of our method using simulations under several scenarios with different values for the transmissibility of each variant, allowing for superspreading and under-reporting as well as differences in natural history between variants (Suppl Sec. 5). The method performed well across all scenarios considered, with a small bias (defined as the difference between the mean posterior estimate and the true *ϵ* value, Fig. 2). MV-EpiEstim was able to accurately estimate the transmission advantage when variants were known to differ in their natural history (characterised by the serial interval distribution, i.e. the delay between onset of symptoms in a case and their infector, Fig. 2c and e). We also explored a scenario typical of real-time outbreak analysis where the natural history of the new variant is different, but in the absence of information, is assumed to be the same as that of the reference.

**Figure 2:**
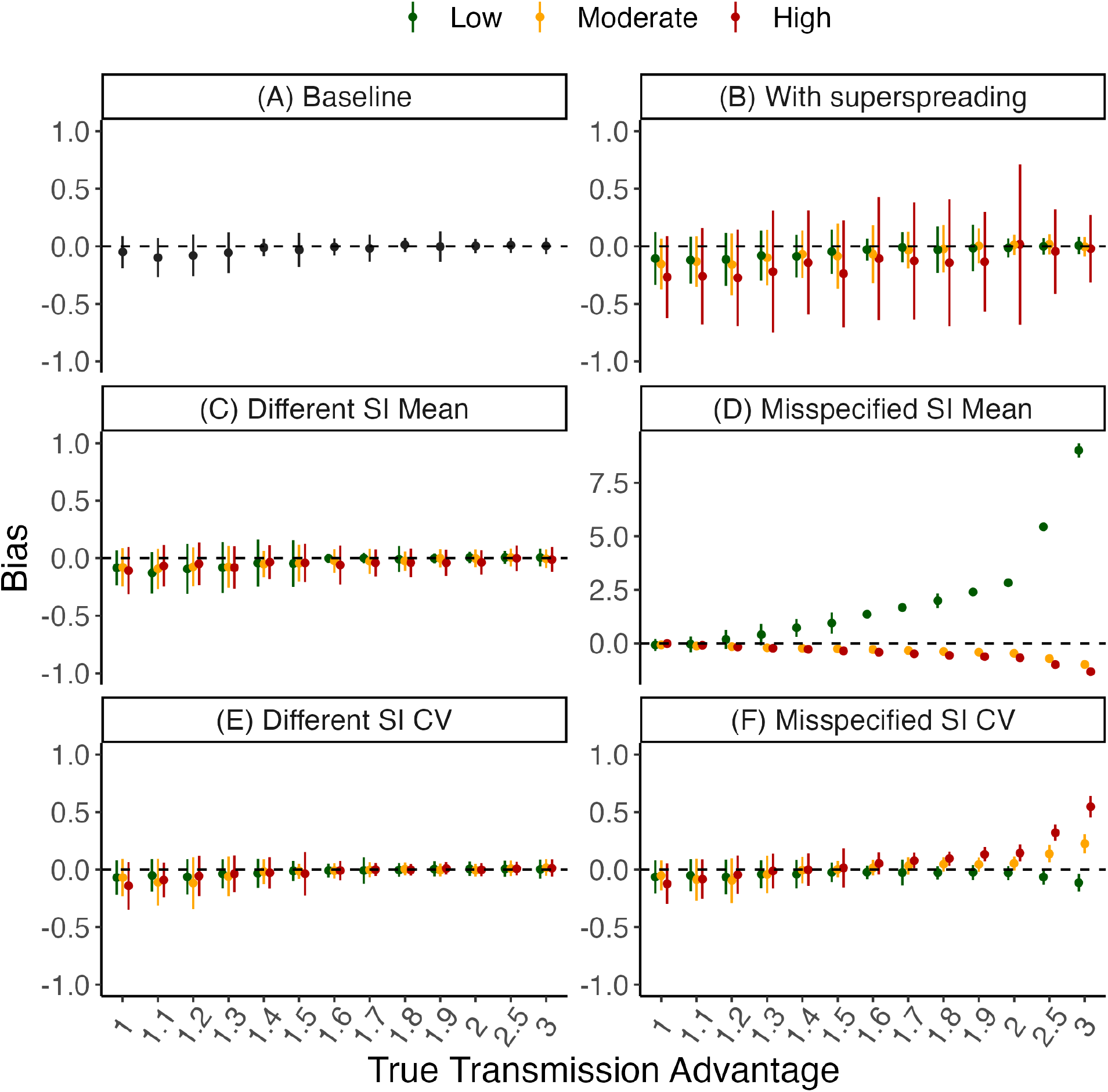
Method performance on simulated data. We assessed the performance of MV-EpiEstim on a range of scenarios. In each panel, the x-axis shows the true value of the effective transmission advantage, *ϵ* (on categorical scale). The y-axis shows the bias i.e., the difference between the posterior mean estimate of the transmission advantage and the true value. The solid dots represent the mean bias (across 100 simulations) and the vertical bars show the standard deviation (SD) of the bias. Each panel corresponds to a different simulation scenario. In all scenarios, the *R*_*t*_ for the reference variant was 1.1 and the *R*_*t*_ for the new variant was *ϵ* times the reference *R*_*t*_ (see Suppl Sec. 5 for details). (A) In the baseline scenario, we assumed no superspreading and the same natural history for both variants. (B) As (A), but with low (overdispersion parameter *k* = 1), moderate (*k* = 0.5) and high (*k* = 0.1) levels of superspreading. (C) As (A), but the mean serial interval of the new variant is 0.5 (low), 1.5 (moderate) or 2 (high) times that of the reference and is correctly specified during estimation. (D) As (C), but the mean serial interval of the variant is assumed to be the same as that of the reference during estimation. (E) As (A), but the coefficient of variation (CV, ratio of standard deviation to mean) of the serial interval of the new variant is 0.5 (low), 1.5 (moderate) or 2 (high) times that of the reference and is correctly specified during estimation. (F) As (E), but the CV of the serial interval of the new variant is assumed to be the same as that of the reference during estimation. Note that the y-axis range is different for panel D. Results using *R*_*t*_ = 1.6 for the reference variant and using fewer days of data are presented in Suppl Secs. 5.3 to 5.9. Results using time-varying reference *R*_*t*_ in one or two locations are shown in Suppl Sec. 5.10 and Suppl Sec. 5.11.

Misspecifying the mean serial interval led to substantial biases, especially when the transmission advantage was moderate (more than 1.5) and the mean serial interval of the new variant was much shorter than (less than half) that of the reference (Fig. 2d). Misspecifying the coefficient of variation of the serial interval had little impact on the quality of the estimates, unless the transmission advantage was very high (more than 2, Fig. 2f).

Even in the presence of substantial superspreading (equivalent to that of SARS-CoV-1, Fig. 2b) or poor case-reporting (up to 80% cases not reported, Fig. S18), neither of which is explicitly accounted for by MV-EpiEstim, the transmission advantage remained unbiased.

In all scenarios, using more days of data reduced both the bias and the uncertainty in the estimated effective transmission advantage (Suppl Secs. 5.3 to 5.7).

We used the full posterior distribution of *ϵ* to classify the variant as more or less transmissible than the reference (see Methods). Crucially, in many scenarios including some where the bias was large, MV-EpiEstim was able to correctly characterise a variant as being more transmissible than the reference. For instance, when the mean serial interval of the new variant was shorter but misspecified, the variant was still correctly classified as more transmissible since *ϵ* was over-estimated (Fig. S10, scenario type low). Conversely, when the mean serial interval of the new variant was longer but was misspecified, correct classification was only feasible with sufficient days of data and a large transmission advantage (Fig. S10, scenario type high).

More results using fewer days of data, two locations, time-varying *R*_*t*_ and accounting for under-reporting are shown in Suppl Sec. 5.

## Discussion

In this study we present a novel method, MV-EpiEstim, to estimate the transmission advantage of a new variant of a pathogen over a reference variant. MV-EpiEstim builds on the EpiEstim method [17], which was found to perform better than other approaches for estimating the instantaneous reproduction number [23]. As such, MV-EpiEstim offers the same functionalities as EpiEstim, including explicitly accounting for imported cases [18]. Because MV-EpiEstim is based on analytical formulations of the marginal posterior densities, the run time of a typical analysis is very short (a few minutes at most on a standard laptop for all analyses presented here). MV-EpiEstim is implemented as a new function (“estimate advantage”) in the R package EpiEstim [24].

We show that MV-EpiEstim could have precisely estimated the effective transmission advantage of the SARS-CoV-2 Alpha variant a few weeks before the earliest published estimate. Importantly, our method only requires as inputs time-series of incident cases and serial interval distributions for each variant. If specific bio-markers are sufficient to distinguish variants (e.g. S-gene), no Whole-Genome-Sequence data is required. Therefore, MV-EpiEstim could be used in near real-time, relying only on routinely collected incidence data and not necessarily suffering from potential delays in the sequencing pipeline.

Our method works well across a range of simulated scenarios, designed to mimic a variety of real-time epidemic contexts, including in the presence of superspreading and when the natural history of the new variant is imperfectly characterised. In the absence of precise information on this natural history, the fast run time offers the possibility of exploring various assumptions and in turn estimate a range of plausible transmission advantages. Our method is robust to under-reporting and temporal changes in reporting if these affect both the reference and the variant equally.

Importantly, we show that our method can accurately characterise a variant as being ‘more’ or ‘less’ transmissible than a reference variant. This simple but robust characterisation could be as important as estimating the exact value of the transmission advantage, especially in informing public health response during the early emergence of a new variant.

We emphasise that our method estimates the *effective* transmission advantage, which will often reflect a combination of several factors such as a true increase in underlying transmissibility and the ability of a new variant to escape immunity. Disentangling these effects is particularly challenging in the context of changing population immunity e.g. due to vaccination roll-out, and may require additional data [25]. However, regardless of its drivers, early identification of a transmission advantage is a critical first step to a timely response.

MV-EpiEstim allows combining information across time and locations, assuming that the effective transmission advantage is constant across these. This allows reducing the uncertainty in the estimates. Temporal or spatial heterogeneity in the transmission advantage (e.g. reflecting heterogeneity in population immunity) can also be characterised by applying the method separately by location or time period, which is easy to do in our software.

Our estimated transmission advantage of the SARS-CoV-2 Alpha variant (over the wildtype) is consistent with those from bespoke analyses using multiple data streams including whole-genome-sequence data [26–30].

The incidence of SARS-CoV-2 in much of the world is still high with nearly 3 million cases reported every week in November 2021 [31]. Given the continued high levels of SARS-CoV-2 transmission and low vaccination coverage globally [32], new variants are likely to continue emerging. Our tool can be used to monitor their transmissibility and rapidly identify variants of concern.

Applications of our work are not limited to SARS-CoV-2; our generic method could easily be used to monitor other pathogens with multiple co-circulating strains such as influenza or *streptococcus pneumoniae*.

## Data Availability

All data used in the study are available online at https://github.com/mrc-ide/epiestims

## Methods

We extend the methodology from Cori et al. [17] and Thompson et al. [18] to develop an inference framework for jointly estimating the transmissibility (instantaneous reproduction number *R*_*t*_) of a reference variant and the effective transmission advantage of novel variants, compared to the reference. For simplicity, we present the method for two variants only (a reference and a new variant). The method is applicable to, and has been implemented for, estimating the transmission advantages of multiple variants over a single reference.

### Assumptions

Our method relies on daily incidence data of the reference and the variant. Where data from more than one location are used, we assume that the epidemic in each location are independent and closed. That is, we do not account for spatial interaction between various locations and assume that all new cases in any location arise from previously infected cases in that location unless identified as imported cases in the dataset. The effective reproduction number is defined as the ratio of locally infected cases to the total infectiousness (due to local or imported cases) in a location. For more details, see [18].

### Notations

We use the following notations:

- Indexes *t* for time, *l* for location and *v* for variant, with *v* = 0 denoting the reference variant and *v* = 1 the new variant,
- *n*_*l*_ the number of locations considered
- *T* the number of days of observation
- 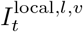 denotes the number of locally infected incident cases of variant *v* at time *t* in location *l*,
- 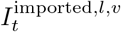 denotes the number of imported infected incident cases of variant *v* at time *t* in location *l* (in the absence of information on imported cases, 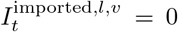 except on the first day of observation, where all cases are assumed to be imported),
- 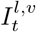 denotes the total number of incident cases of variant *v* at time *t* in location *l*, with 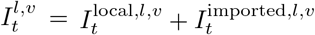,
- 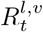 denotes the instantaneous reproduction number for variant *v* at time *t* in location *l*. For simplicity we use 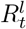 to denote the instantaneous reproduction number for the reference variant in location *l* i.e.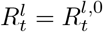.
- *w*^*v*^ is the probability mass function of the discrete serial interval for variant *v*, assumed the same across all locations, but potentially different between variants (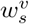 is the probability that the serial interval lasts s days, *s* = 1, …, *SI*_*max*_; and we assume 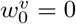).
- 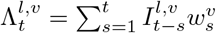 is the overall infectiousness for variant *v* at time *t* and in location *l* due to past incident cases of that variant in that location (both imported and locally infected cases).
- For simplicity we introduce the generic notation 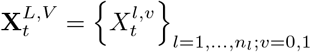 for the variable *X* at time *t* across all locations and both variants.

We assume that 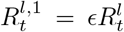, i.e. the reproduction number of the new variant is proportional to that for the reference variant at all times and in all locations; the proportional factor *ϵ* is the effective transmission advantage (if *ϵ >* 1, or disadvantage if *ϵ* < 1) of the new variant compared to the reference variant, assumed constant over time and across locations. We explored values of *ϵ >* 1 in all simulation scenarios as values of *ϵ <* 1 correspond to swapping the reference and new variant.

We assume the number of secondary infections generated by each case is Poisson distributed. Under these assumptions, the likelihood of the time series of incident cases of the reference and the new variants can be written as

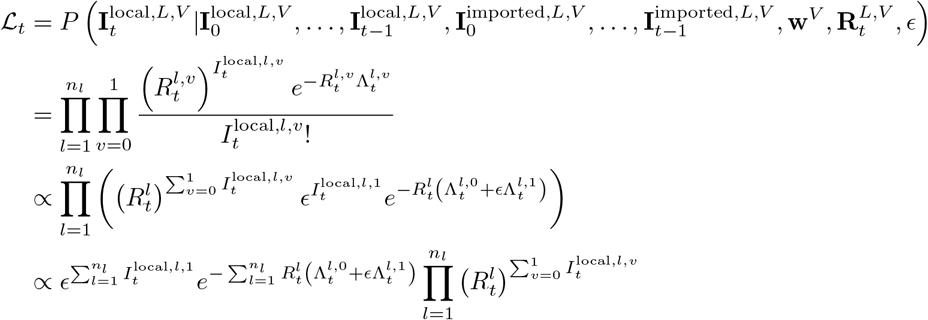

We assume Gamma priors for each 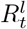, with same shape *a* and scale *b* across times and locations, and for *ϵ*, with shape *c* and scale *d*. The joint posterior distribution of parameters given the observations is (assuming the serial interval distributions for both variants **w**^*V*^ are known):

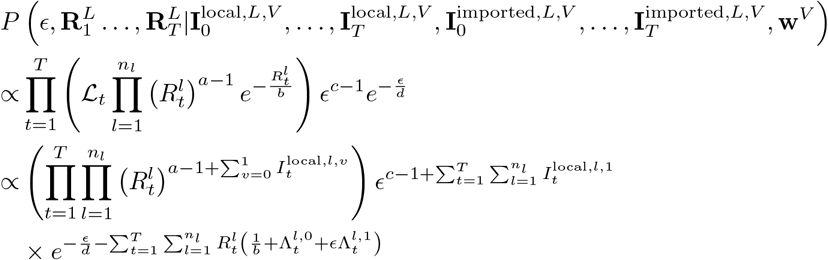

The marginal posterior distribution for *ϵ* given the data (i.e. the incidence for all variants, at all locations and for all time steps) and given the reproduction number for the reference variant in all locations and at all time steps is given by:

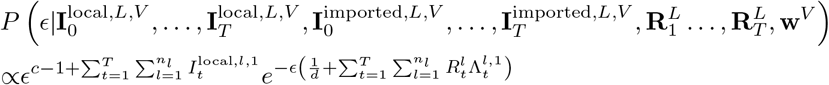

Therefore, the marginal posterior distribution of *ϵ* given the data and other parameters is a Gamma distribution with shape 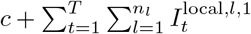 and scale 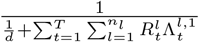.

Similarly, the marginal posterior distribution for 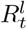 at time step *t* and in location *l* given the data, *ϵ*, and the reproduction number at other locations and time steps, is given by:

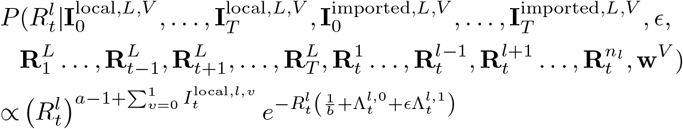

Therefore, the marginal posterior distribution of 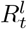 given the data and other parameters is a Gamma distribution with shape 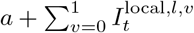 and scale 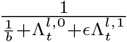.

### Monte Carlo Markov Chain (MCMC) inference

The analytical formulation of the marginal posterior distributions for 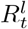 and *ϵ* allow us to use a multi-stage Gibbs sampler for the MCMC inference.

To initialize 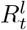, we use EpiEstim to estimate a single reproduction number for the reference variant over the entire time period of observations, and using incidence aggregated across all locations. The posterior mean is then used as the initial value for 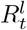. We independently use the same approach to estimate a single reproduction number for the new variant; *ϵ* is then initialised to the median of the ratio of the reproduction numbers for the new variant and the reference.

We first sample from the marginal distribution of 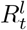, conditional on *ϵ*, and then we sample from the marginal distribution of *ϵ*, conditional on the newly sampled value of 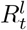. We repeat this procedure for a fixed number of iterations or until convergence is achieved. Convergence is assessed using Gelman-Rubin convergence diagnostic [34] using 1.1 as a cut-off value.

### Choosing a time-period for estimation of *ϵ*

Users can set the time period over which estimation will be carried out. We recommend that the estimation is started after at least one generation of cases has been observed. The default starting point in the software is set to the first day of non-zero incidence across all locations plus the 95^th^ percentile of the serial interval distribution.

### Classification of a variant

We used the posterior distribution of the effective transmission advantage to classify a new variant (in relation to the reference variant) as:

- ‘More transmissible’ if the 2.5^th^ quantile of the posterior distribution was greater than 1;
- ‘Less transmissible’ if the 97.5^th^ quantile of the posterior distribution was less than 1; and,
- ‘Unclear’ if the 95% CrI contained 1.

### Implementation

The inference method is implemented in a new function “estimate advantage” of the development version of the R package EpiEstim available at https://github.com/mrc-ide/EpiEstim.

## Acknowledgements

The use of pillar-2 PCR testing data was made possible thanks to PHE colleagues, and we extend our thanks to N Gent for facilitation and insights into these data. We also thank Edward S Knock for his inputs on the data for England. This study is partially funded by the National Institute for Health Research (NIHR) Health Protection Research Unit in Modelling and Health Economics, a partnership between Public Health England, Imperial College London and LSHTM (grant code NIHR200908); the authors acknowledge funding from the MRC Centre for Global Infectious Disease Analysis (reference MR/R015600/1), which is jointly funded by the UK Medical Research Council (MRC) and the UK Foreign, Commonwealth & Development Office (FCDO), under the MRC/FCDO Concordat agreement and is also part of the EDCTP2 programme supported by the European Union. JW acknowledges research funding from the Wellcome Trust (grant 102169/Z/13/Z). SB acknowledges funding from the Wellcome Trust (grant 219415). RKN acknowledges funding from the Medical Research Council Doctoral Training Partnership. Disclaimer: The views expressed are those of the author(s) and not necessarily those of the NIHR, Public Health England or the Department of Health and Social Care.

