## Supplementary Information for "A generic method and software to estimate the transmission advantage of pathogen variants in real-time : SARS-CoV-2 as a case-study"

**6 Code and Data availability** **34**

#### 1 Overview

In this document, we present details of the SARS-CoV-2 variant-specific incidence data used in the analysis (Sec. 2) and describe the method used for obtaining estimates of the transmission advantage for SARS-CoV-2, both using a naive approach and using MV-EpiEstim (Sec. 3). Sec. 4 shows additional results for the estimation of the transmission advantage of SARS-CoV-2 variants of concern (VOCs), including more detailed results on the estimated transmission advantage of Alpha over the wildtype and estimates of the transmission advantages of Beta and Gamma (combined) over the wildtype. Sec. 5 presents an overview of the simulation study used to assess the validity of our method. We describe the methodology used for the simulation and present the range of scenarios we explored, as well as more comprehensive results from our simulation study.

#### 2 SARS-CoV-2 variant-specific incidence data

##### 2.1 Incidence data from England

We used the daily number of positive tests from England's community SARS-CoV-2 testing system (also called Pillar 2) from 1st September 2020 to 20th June 2021, stratified by NHS region (Fig S1). The Pillar 2 testing data were shared with Imperial College London by Public Health England. Up to 14th March 2021, we interpreted the number of samples with no S-gene target failure in this data as incidence of the wildtype. Samples with S-gene failure were considered to be of the Alpha variant throughout.

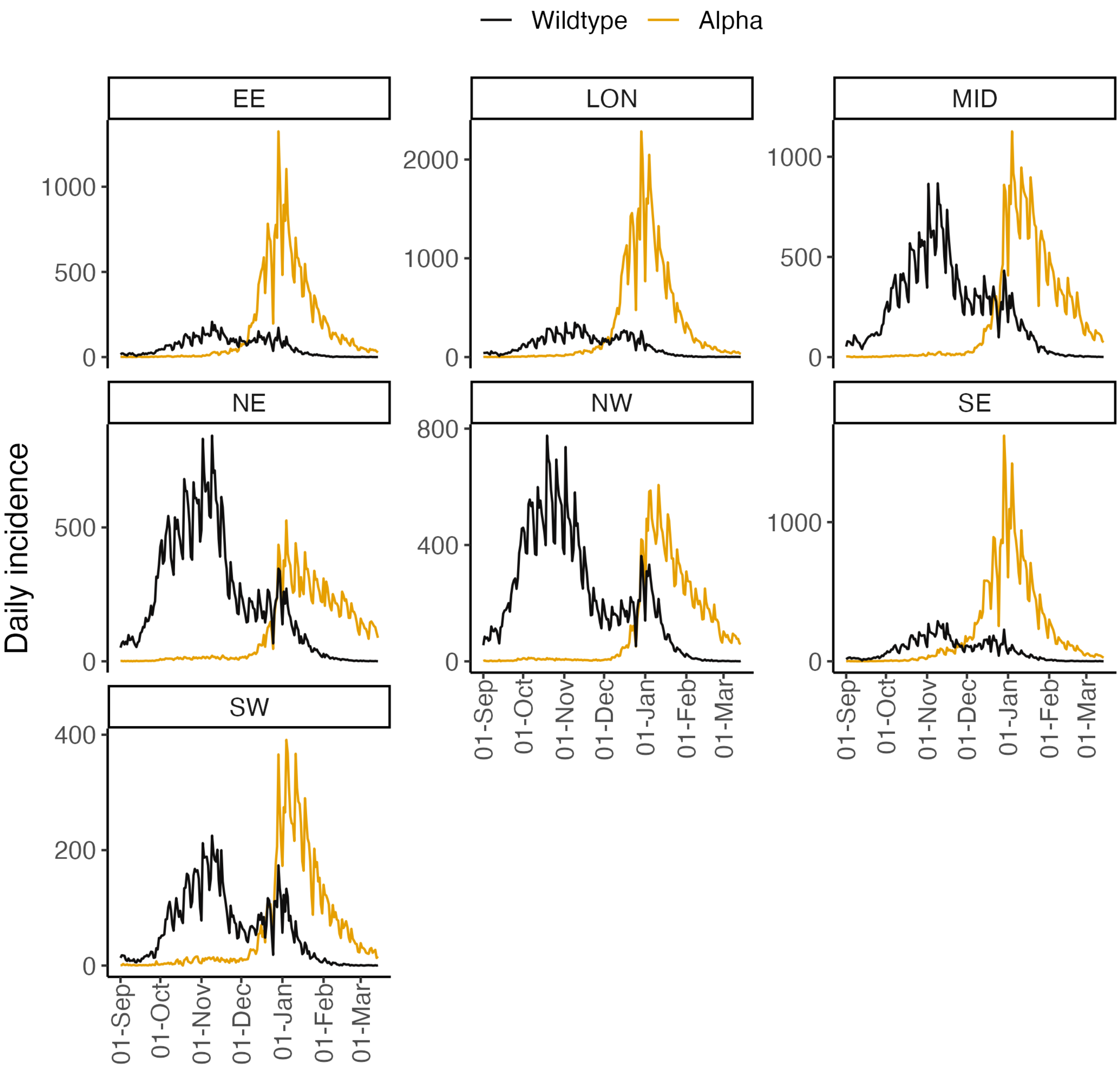

Figure S1: Daily reported incidence of SARS-CoV-2 wildtype (black) and Alpha (orange) variants in the 7 NHS regions in England. Note that the y-axis in each panel is different. The NHS England regions are - East of England (EE), London (LON), Midlands (MID), North-East (NE), North-West (NW), South-East (SE), South-West (SW).

##### 2.2 Incidence data from France

Santé Publique France reports the age-disaggregated number of PCR tests with Alpha, Beta, and Gamma variants of SARS-CoV-2 at a sub-national level in France [1] with the incidence of Beta and Gamma variants reported as an aggregate. The absence of labelling with a specific VOC was interpreted as an infection with the wildtype. The 18 ADM2 units for which data were reported include metropolitan France and overseas regions. We aggregated the data across all age groups to obtain a daily incidence time series for each variant from 28$^{th}$ February to 30$^{th}$ May 2021 (Fig S2).

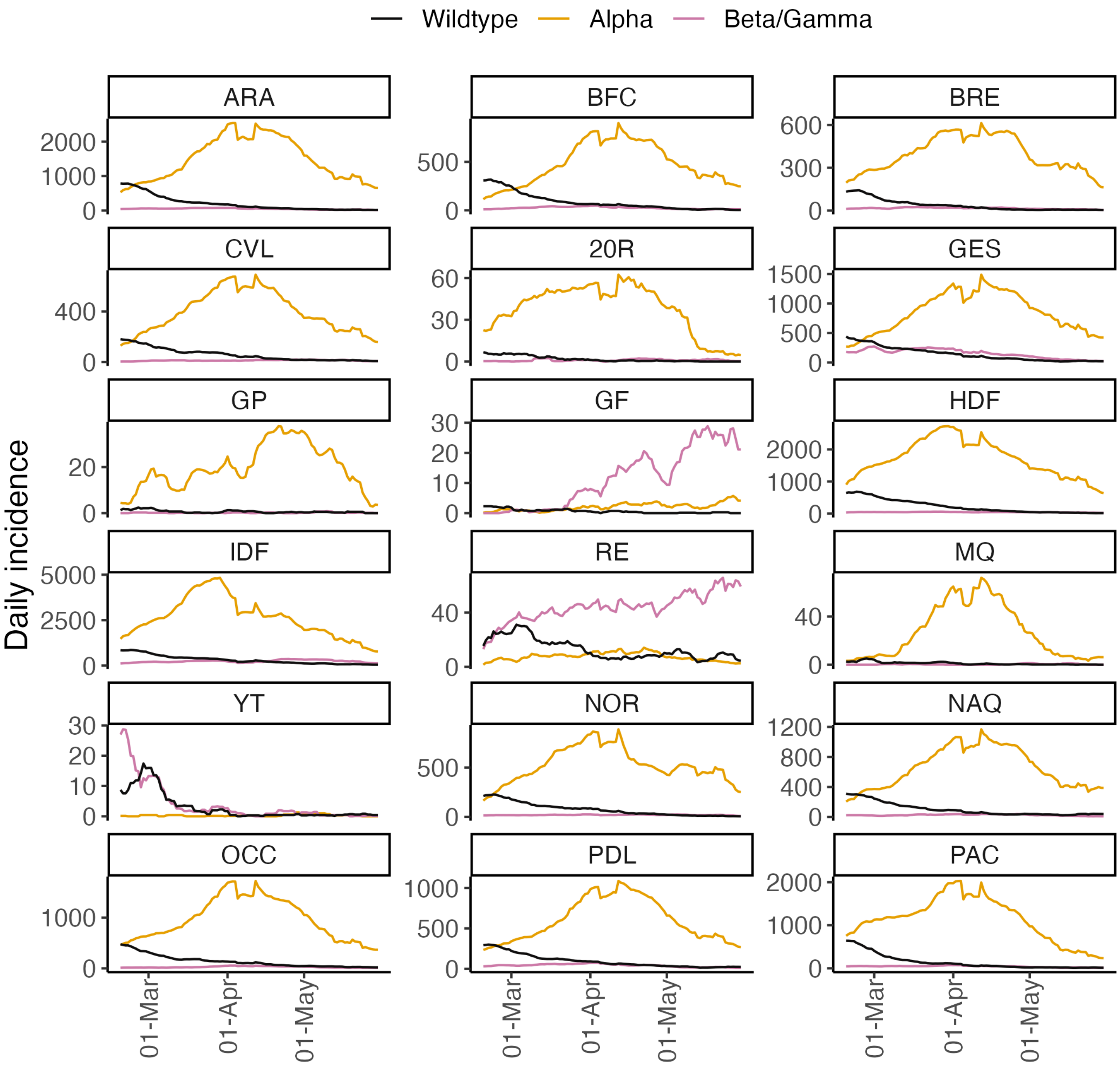

Figure S2: Daily incidence of SARS-CoV-2 wildtype (black), Alpha (orange) and Beta/Gamma (blue) variants for the 18 ADM2 regions in France. Note that the y-axis in each panel is different. The ADM2 regions are - ARA : Auvergne-Rhône-Alpes, BFC : Bourgogne-Franche-Comté, BRE : Bretagne, CVL : Centre-Val de Loire, 20R : Corse, GES : Grand Est, GP : Guadeloupe, GF : Guyane, HDF : Hauts-de-France, IDF : Île-de-France, RE : La Réunion, MQ : Martinique, YT : Mayotte, NOR : Normandie, NAQ : Nouvelle-Aquitaine, OCC : Occitanie, PDL : Pays de la Loire, and PAC : Provence-Alpes-Côte d'Azur.

### 3 Estimating the effective transmission advantage

#### 3.1 Serial interval distribution

Both the naive (see next paragraph) and the MV-EpiEstim approaches use the discrete distribution of the serial interval (time between symptom onset in a case and their infector) as an input. We assumed a discrete gamma distributed serial interval for SARS-CoV-2 with mean 5.4 days and standard deviation of 1.5 days following [2]. We used the same serial interval distribution across all variants.

##### 3.2 Naive approach

To obtain a naive estimate of the effective transmission advantage of a VOC over the reference SARS-CoV-2 variant, we first estimated the daily effective reproduction number independently for each variant (wildtype or VOC) for 18 ADM2 units in France and 7 NHS regions in England using the R package EpiEstim [3]. We used a sliding weekly window, and set the prior $R_t$ to have a mean and a standard deviation of 1.

To exclude region-weeks where the $R_t$ estimates were highly uncertain, we only used estimates from region-weeks where the width of 95% CrI of $R_t$ was less than 0.5. We started estimation of $R_t$ on the week starting on the $11^{\text{th}}$ day. The threshold of 11 days was chosen as it is the $99^{\text{th}}$ percentile of the serial interval distribution. That is, 99% of the cases that were infected by an index cases from day 1 in our analysis are expected to have been observed by day 11. Note that because of these exclusion criteria, some of the naive estimates are missing in the tables shown in section Sec. 4, when no weeks could be included for a particular region or time period.

For each region-week included in the analysis, we drew a sample of 100 values from the posterior distribution of $R_t$ for each variant. Naive estimates of a variant's transmission advantage over a reference variant were obtained by dividing the sampled values from their respective posterior $R_t$ distributions (with random pairing). To account for sub-national variation in $R_t$ profile, estimates at the national level were obtained by pooling the sub-national estimates (thereby giving the same weight to each week of data from any region). To gain insight into the potential temporal heterogeneity of the effective transmission advantage, we divided the incidence time series into four non-overlapping periods of equal duration and estimated the transmission advantage in each period.

##### 3.3 Estimation using MV-EpiEstim

We set the priors for both $R_t$ and $\epsilon$ to have mean and standard deviation 1. We ran the multi-stage Gibbs sampler for 20,000 iterations. The first 5,000 iterations were discarded as burn-in and thinning was set to keep 1 in 10 iterations, leading to a final posterior sample of size 1,500.

Posterior samples of the transmission advantage were obtained for (i) each region independently and (ii) nationally but using regional data, by assuming a single underlying transmission advantage and region-specific $R_t$ profiles. Independent estimates of the transmission advantage were obtained for the same non-overlapping time period as for the naive estimates.

To mimic real-time epidemic context and examine how estimates changed as more data became available, we estimated the effective transmission advantage using data available up to successive weeks.

#### 4 Estimates of the effective transmission advantages of SARS-CoV-2 variants

##### 4.1 Alpha over wildtype

###### 4.1.1 England

Main results for the estimated transmission advantage of Alpha over the wildtype using data from England is shown in the main text Figure 1.

| Region/Time Period | Naive | MV-EpiEstim |
|---|---|---|
| All | 1.41 (0.86, 2.01) | 1.46 (1.44, 1.47) |
| East of England | 1.38 (1.02, 2.13) | 1.43 (1.39, 1.47) |
| London | 1.40 (0.93, 1.82) | 1.46 (1.43, 1.50) |
| Midlands | 1.44 (0.92, 2.04) | 1.54 (1.50, 1.58) |
| North East and Yorkshire | 1.41 (0.86, 2.05) | 1.46 (1.42, 1.50) |
| North West | 1.50 (0.71, 2.08) | 1.51 (1.47, 1.55) |
| South East | 1.35 (1.00, 1.87) | 1.36 (1.33, 1.39) |
| South West | 1.41 (0.78, 1.82) | 1.40 (1.35, 1.45) |
| Quarter 1 | 0.89 (0.62, 1.17) | 1.03 (0.99, 1.08) |
| Quarter 2 | 1.36 (0.82, 1.95) | 1.48 (1.45, 1.51) |
| Quarter 3 | 1.45 (1.07, 2.02) | 1.50 (1.48, 1.52) |
| Quarter 4 | 1.39 (0.93, 2.12) | 1.28 (1.23, 1.33) |

Table S1: Estimates of the effective transmission advantage of SARS-CoV-2 Alpha variant over the wildtype using the naive approach and MV-EpiEstim for 7 NHS regions in England and 4 non-overlapping time periods. Estimates shown are the posterior median with 95% CrI in parenthesis. Quarters correspond to - Quarter 1: 11$^{th}$ September - 27$^{th}$ October 2020; Quarter 2: 27$^{th}$ October - 12$^{th}$ December 2020; Quarter 3: 12$^{th}$ December 2020 - 27$^{th}$ January 2021; 27$^{th}$ January - 14$^{th}$ March 2020.

##### 4.1.2 France

| Region/Time Period | Naive | MV-EpiEstim |
|---|---|---|
| All | 1.21 (0.75, 1.65) | 1.29 (1.29, 1.30) |
| Auvergne-Rhône-Alpes | 1.25 (0.89, 1.67) | 1.40 (1.38, 1.43) |
| Bourgogne-Franche-Comté | 1.27 (0.90, 1.78) | 1.41 (1.37, 1.46) |
| Bretagne | 1.34 (0.89, 1.78) | 1.35 (1.29, 1.40) |
| Centre-Val de Loire | 1.18 (0.79, 1.61) | 1.32 (1.27, 1.36) |
| Corse | - | 1.15 (1.00, 1.31) |
| Grand Est | 1.25 (0.70, 1.48) | 1.31 (1.28, 1.34) |
| Guadeloupe | - | 1.12 (0.95, 1.35) |
| Guyane | - | 1.33 (1.05, 1.66) |
| Hauts-de-France | 1.19 (0.97, 1.41) | 1.28 (1.26, 1.30) |
| Île-de-France | 1.08 (0.87, 1.47) | 1.21 (1.20, 1.23) |
| La Réunion | 1.14 (0.67, 2.08) | 1.07 (0.98, 1.18) |
| Martinique | - | 1.27 (1.09, 1.49) |
| Mayotte | - | 1.13 (0.73, 1.69) |
| Normandie | 1.21 (0.91, 1.59) | 1.31 (1.28, 1.35) |
| Nouvelle-Aquitaine | 1.22 (0.70, 1.62) | 1.28 (1.25, 1.31) |
| Occitanie | 1.18 (0.81, 1.50) | 1.27 (1.25, 1.30) |
| Pays de la Loire | 1.20 (0.66, 1.55) | 1.29 (1.26, 1.32) |
| Provence-Alpes-Côte d'Azur | 1.22 (0.67, 1.58) | 1.37 (1.34, 1.40) |
| Quarter 1 | 1.45 (1.26, 1.71) | 1.42 (1.41, 1.44) |
| Quarter 2 | 1.28 (0.99, 1.62) | 1.24 (1.22, 1.25) |
| Quarter 3 | 1.15 (0.83, 1.51) | 1.11 (1.09, 1.13) |
| Quarter 4 | 1.00 (0.67, 1.52) | 0.97 (0.95, 0.99) |

Table S2: Estimates of the effective transmission advantage of SARS-CoV-2 Alpha variant over the wildtype using the naive approach and MV-EpiEstim for 18 ADM2 regions in France and 4 non-overlapping time periods. Estimates shown are the posterior median with 95% CrI in parenthesis. Quarters correspond to - Quarter 1: 28$^{th}$ February - 23$^{rd}$ March 2021; Quarter 2: 23$^{rd}$ March - 14$^{th}$ April 2021; Quarter 3: 14$^{th}$ April - 7$^{th}$ May 2021; 7$^{th}$ May - 20$^{th}$ May 2021.

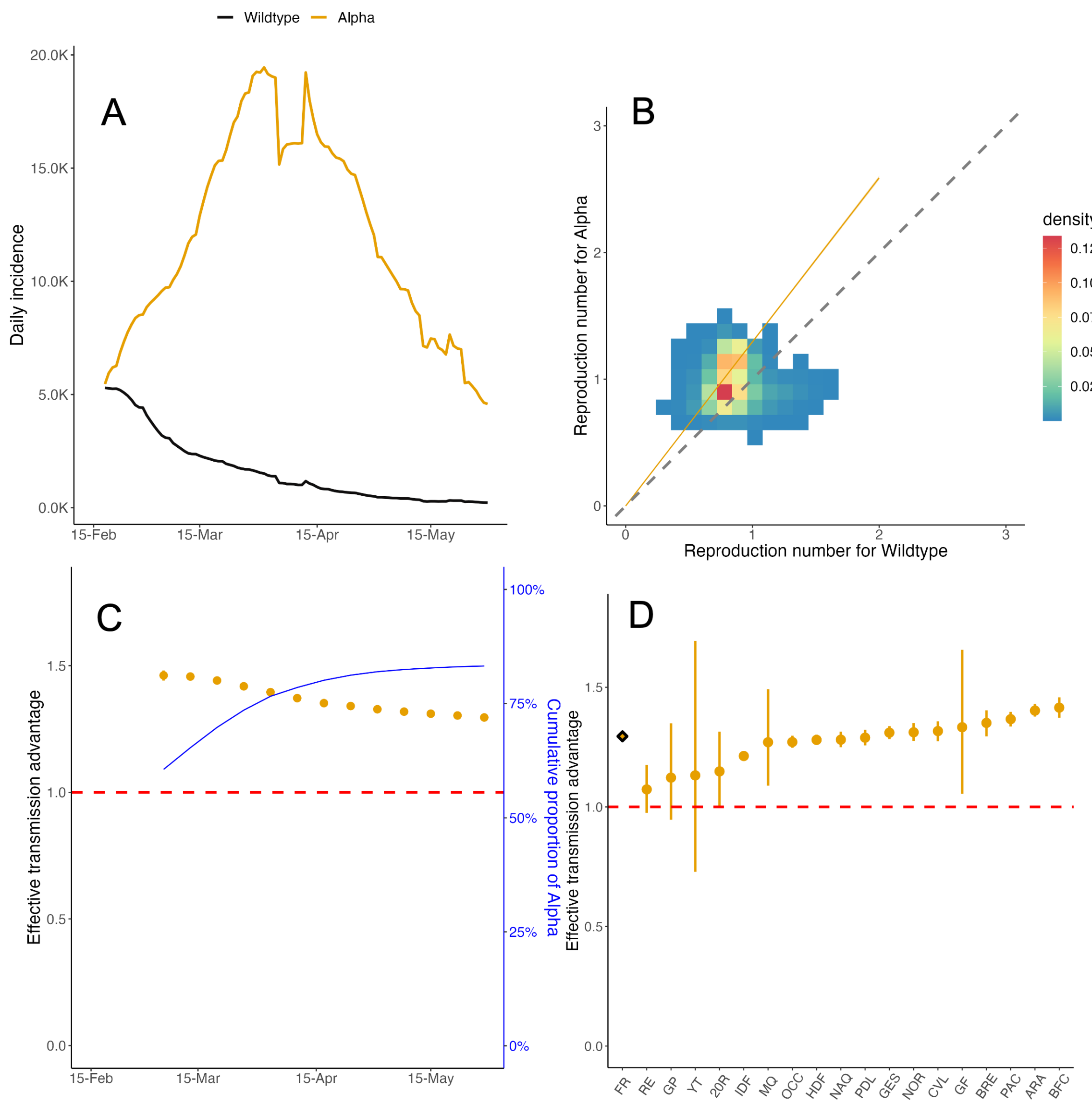

Figure S3: **Effective transmission advantage of Alpha over wildtype in France** (A) The daily reported incidence of cases of the wildtype (black) and Alpha (orange) in France from 18$^{th}$ February to 30$^{th}$ May 2021. (B) The effective reproduction number $R_t$ estimated independently for the wildtype (x-axis) and Alpha (y-axis) on sliding weekly windows. The color of the cells indicates the density of the draws from the respective posterior distributions of $R_t$. The dashed diagonal line indicates the $x = y$ threshold. Coloured cells lying above the diagonal line suggest that Alpha is more transmissible. The orange line denotes the median effective transmission advantage estimated using MV-EpiEstim. 95% CrI were so narrow that they could not be distinguished from the line. (C) Effective transmission advantage estimated from MV-EpiEstim using data available up to the date specified on the x-axis. The dark blue line denotes the proportion of cumulative incidence of Alpha (right y-axis) counted from 18$^{st}$ February 2021. (D) Effective transmission advantage estimated using MV-EpiEstim for all ADM2 region in France together (diamond) and separately (solid circle) using data from 18$^{th}$ February to 30$^{th}$ May 2021. The ADM2 regions are - ARA : Auvergne-Rhône-Alpes, BFC : Bourgogne-Franche-Comté, BRE : Bretagne, CVL : Centre-Val de Loire, 20R : Corse, GES : Grand Est, GP : Guadeloupe, GF : Guyane, HDF : Hauts-de-France, IDF : Île-de-France, RE : La Réunion, MQ : Martinique, YT : Mayotte, NOR : Normandie, NAQ : Nouvelle-Aquitaine, OCC : Occitanie, PDL : Pays de la Loire, and PAC : Provence-Alpes-Côte d'Azur. In panels (C) and (D), the solid circles denote the median estimate, the vertical lines indicate the 95% CrI, and the red dashed line denotes the $\epsilon = 1$ threshold.

#### 4.2 Beta and Gamma over wildtype (France)

| Region/Time-period | Naive | MV-EpiEstim |
|---|---|---|
| All | 1.17 (0.69, 1.74) | 1.25 (1.24, 1.27) |
| Auvergne-Rhône-Alpes | 1.15 (0.88, 1.50) | 1.29 (1.25, 1.34) |
| Bourgogne-Franche-Comté | 1.22 (0.77, 1.90) | 1.32 (1.25, 1.38) |
| Bretagne | 1.26 (0.82, 1.89) | 1.28 (1.20, 1.37) |
| Centre-Val de Loire | 1.15 (0.68, 1.73) | 1.35 (1.27, 1.44) |
| Corse | - | 1.30 (1.03, 1.62) |
| Grand Est | 1.08 (0.61, 1.31) | 1.15 (1.12, 1.17) |
| Guadeloupe | - | 1.07 (0.61, 1.67) |
| Guyane | - | 1.40 (1.14, 1.71) |
| Hauts-de-France | 1.21 (0.96, 1.46) | 1.26 (1.22, 1.31) |
| Île-de-France | 1.16 (0.85, 1.72) | 1.27 (1.25, 1.29) |
| La Réunion | 1.32 (0.89, 2.10) | 1.15 (1.07, 1.22) |
| Martinique | - | 1.30 (0.90, 1.84) |
| Mayotte | 0.95 (0.47, 1.54) | 0.83 (0.69, 1.00) |
| Normandie | 1.20 (0.88, 1.70) | 1.28 (1.22, 1.35) |
| Nouvelle-Aquitaine | 1.16 (0.53, 1.67) | 1.23 (1.17, 1.29) |
| Occitanie | 1.16 (0.71, 1.77) | 1.29 (1.24, 1.35) |
| Pays de la Loire | 1.12 (0.64, 1.65) | 1.19 (1.14, 1.24) |
| Provence-Alpes-Côte d'Azur | 1.21 (0.65, 1.63) | 1.33 (1.28, 1.38) |
| Quarter 1 | 1.31 (0.85, 1.78) | 1.28 (1.26, 1.30) |
| Quarter 2 | 1.17 (0.87, 1.69) | 1.15 (1.13, 1.17) |
| Quarter 3 | 1.17 (0.81, 1.73) | 1.20 (1.17, 1.23) |
| Quarter 4 | 1.01 (0.58, 1.75) | 0.97 (0.94, 0.99) |

Table S3: Estimates of the combined effective transmission advantage of SARS-CoV-2 Beta and Gamma variants over the wildtype in France using the naive approach and MV-EpiEstim for 18 ADM2 regions in France and 4 non-overlapping time periods. Estimates shown are the posterior median with 95% CrI in parenthesis. Quarters correspond to - Quarter 1: 28$^{th}$ February - 23$^{rd}$ March 2021; Quarter 2: 23$^{rd}$ March - 14$^{th}$ April 2021; Quarter 3: 14$^{th}$ April - 7$^{th}$ May 2021; 7$^{th}$ May - 20$^{th}$ May 2021.

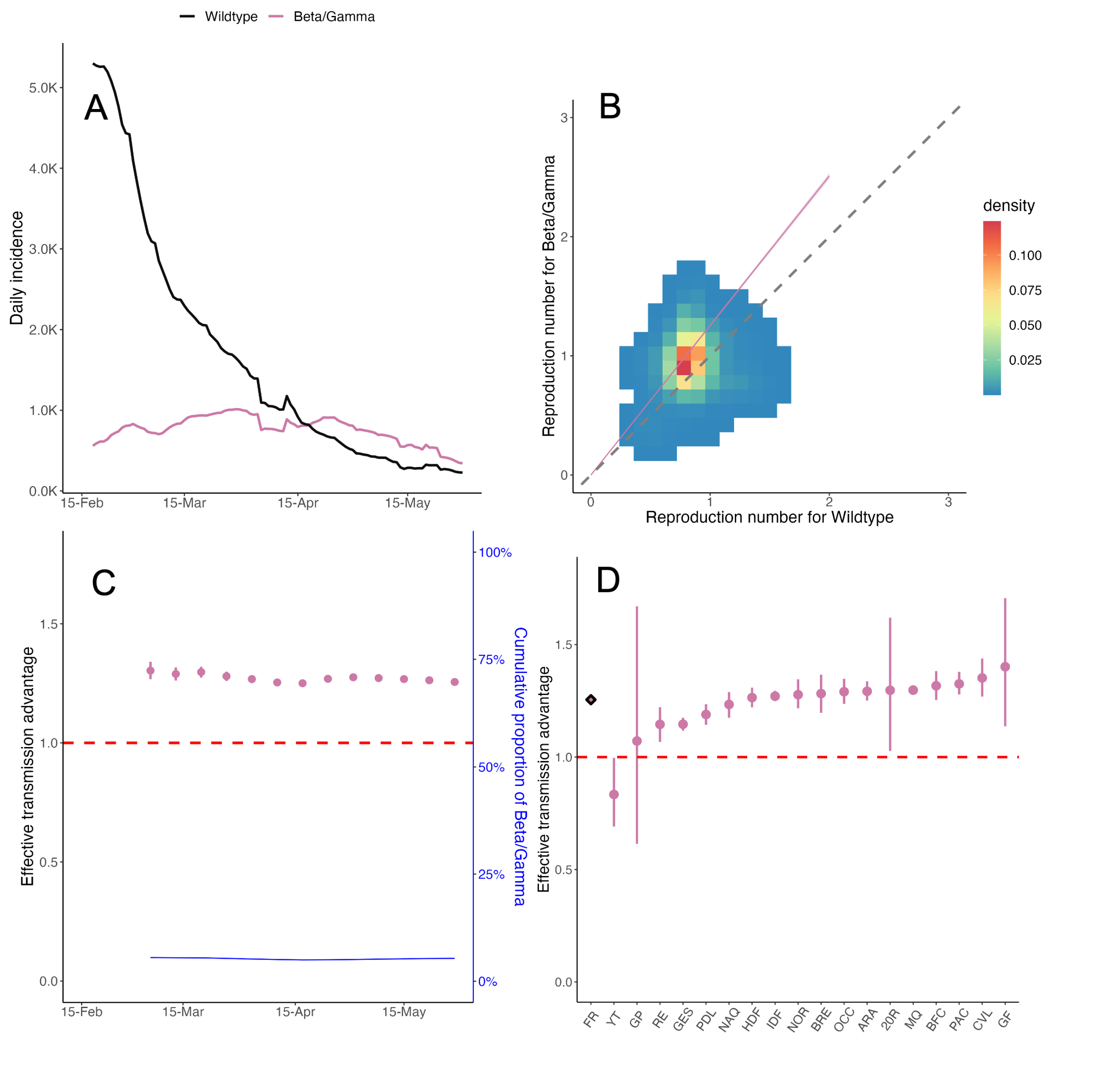

Figure S4: **Effective transmission advantage of Beta and Gamma (combined) over wildtype in France** (A) The daily reported incidence of cases of the wildtype (black) and Beta/Gamma (blue) in France 18$^{th}$ February to 30$^{th}$ May 2021. (B) The effective reproduction number $R_t$ estimated independently for the wildtype (x-axis) and Beta/Gamma (y-axis) on sliding weekly windows. The color of the cells indicates the density of the draws from the respective posterior distributions of $R_t$. The dashed diagonal line indicates the $x = y$ threshold. Coloured cells lying above the diagonal line suggest that Beta/Gamma is more transmissible. The pink line denotes the median effective transmission advantage estimated using MV-EpiEstim. 95% CrI were so narrow that they could not be distinguished from the line. (C) Effective transmission advantage estimated from MV-EpiEstim using data available up to the date specified on the x-axis. The blue line denotes the proportion of cumulative incidence of Beta/Gamma (right y-axis) counted from 18$^{th}$ 2021. (D) Effective transmission advantage estimated using MV-EpiEstim for all ADM2 regions in France together (diamond) and separately (solid circles) using data from 18$^{th}$ February to 30$^{th}$ May 2021. The ADM2 regions are - ARA : Auvergne-Rhône-Alpes, BFC : Bourgogne-Franche-Comté, BRE : Bretagne, CVL : Centre-Val de Loire, 20R : Corse, GES : Grand Est, GP : Guadeloupe, GF : Guyane, HDF : Hauts-de-France, IDF : Île-de-France, RE : La Réunion, MQ : Martinique, YT : Mayotte, NOR : Normandie, NAQ : Nouvelle-Aquitaine, OCC : Occitanie, PDL : Pays de la Loire, and PAC : Provence-Alpes-Côte d'Azur. In panels (C) and (D), the solid circles denote the median estimate, the vertical lines indicate the 95% CrI, and the red dashed line denotes the $\epsilon = 1$ threshold.

### 5 Method performance using simulated data

#### 5.1 Simulation approach

We simulated SARS-CoV-2-like incidence data using a branching process where daily incidence is assumed to follow a Poisson distribution:

$$I_t \sim Poisson(R_t \sum_{s=1}^{t-1} I_s \omega_{t-s}), \quad (1)$$

where $\omega_s$ is the probability mass function of the discrete serial interval.

We assume that the effective reproduction number for the variant is $\epsilon \times R_t$, where $R_t$ is the effective reproduction number for the reference variant and $\epsilon$ is the effective transmission advantage of the new variant over the reference. We explored values of $\epsilon > 1$ in all simulation scenarios as $\epsilon < 1$ corresponds to swapping the reference and new variant.

We seeded the epidemic with 20 cases of the reference variant for 10 successive days and 1 case of the new variant on the $10^{th}$ day. We then simulated forward for an additional 100 days, generating 100 stochastic epidemic trajectories for each simulation scenario and each combination of parameters considered for that scenario (Tab S4). We then estimated the effective transmission advantage using 10, 20, 30, or 50 days of data counted from the $11^{th}$ day (see Sec. 3.2).

In each simulation scenario, we assessed the performance of the method using the following metrics:

- Bias, defined as difference between the mean posterior estimate of the effective transmission advantage and its true value;
- Uncertainty, defined as the posterior standard deviation (SD);
- Coverage probability, defined as the proportion of simulations where the 95% CrI of the transmission advantage contained the true value;
- Classification. We used the posterior distribution of $\epsilon$ to classify the new variant as "more transmissible", "less transmissible" than the reference or "unclear" (see methods in main text). To assess the classification performance, we consider the proportion of simulations where the variant is classified correctly (when the true value of $\epsilon$ is 1, we consider the correct classification to be 'unclear').

| Parameter | Values |
|---|---|
| Reference $R_t$ | 1.1, 1.6 |
| Mean of reference serial interval | 5.4 days |
| Standard deviation of reference serial interval | 1.5 days |
| Mean of variant serial interval | Reference serial interval mean $\times$ (0.5, 1, 1.5, 2) |
| CV of variant serial interval | Reference serial interval CV $\times$ (0.5, 1, 1.5, 2) |
| Overdispersion | 0.1, 0.5, 1 |

Table S4: Parameter values used in the simulations. For each simulation scenario, we considered all (relevant) combinations of parameter values shown in this table; and for each parameter combination, we simulated 100 data sets. CV: coefficient of variation

We first considered a baseline scenario (Sec. 5.3), where we assumed that the natural history of the reference and the new variants are same. We relaxed this assumption in other scenarios, assuming either that the SI distribution of the variant has a different mean (Sec. 5.4) or CV (Sec. 5.6), but that the SI distribution of both the new and reference variants are correctly specified in MV-EpiEstim. We then explored a scenario typical of real-time outbreak analysis where the SI distribution (mean or CV) of the variant is different from that of the reference but in the absence of more information, is assumed to be the same as that of the reference (Secs. 5.5 and 5.7). We also explored the performance of our method in the presence of superspreading (Sec. 5.8), extending the simulation to use a negative binomial offspring distribution, i.e.:

$$I_t \sim NegBin(R_t \sum_{s=1}^{t-1} I_s \omega_{t-s}, \kappa \sum_{s=1}^{t-1} I_s \omega_{t-s}), \quad (2)$$

where $\kappa$ is the overdispersion parameter (lower values of $\kappa$ denoting higher levels of superspreading). Finally, we assessed the sensitivity of MV-EpiEstim to under-reporting (Sec. 5.9), assuming a constant reporting rate for the reference and the new variant.

The scenarios outlined above used a constant $R_t$ over the period of the simulation. We also explored the effect of time-varying $R_t$ profiles on method performance (Sec. 5.10). Finally, we considered simulations with two locations with time-varying $R_t$ (Sec. 5.11), simulating independent epidemics in each location as described above.

#### 5.2 Description of figures

The figures that follow in the remainder of Sec. 5 are composed of four panels. In each figure panel, we present a performance indicator summarised across 100 simulations. In each figure, the top-left panel shows the mean $\pm$ SD of the bias in the estimate of the effective transmission advantage. The dashed horizontal line denotes the threshold bias of 0. The top-right panel shows the mean $\pm$ SD of the uncertainty in estimates. The bottom-left panel shows the coverage probability (mean and 95% binomial confidence interval (CI)). The dashed horizontal line denotes the threshold value of 0.95. The bottom-right panel shows classification performance (mean and 95% binomial CI). For definition of each performance indicator, see Sec. 5.1.

#### 5.3 Baseline scenario

In this section, we present the results for the baseline scenario. That is, we assume no superspreading, and that both the reference and the new variant have the same natural history. Results are shown for estimates obtained using 10, 20, 30 and 50 days of data in MV-EpiEstim.

(A) $R_t$ = 1.1 and 10 days of data

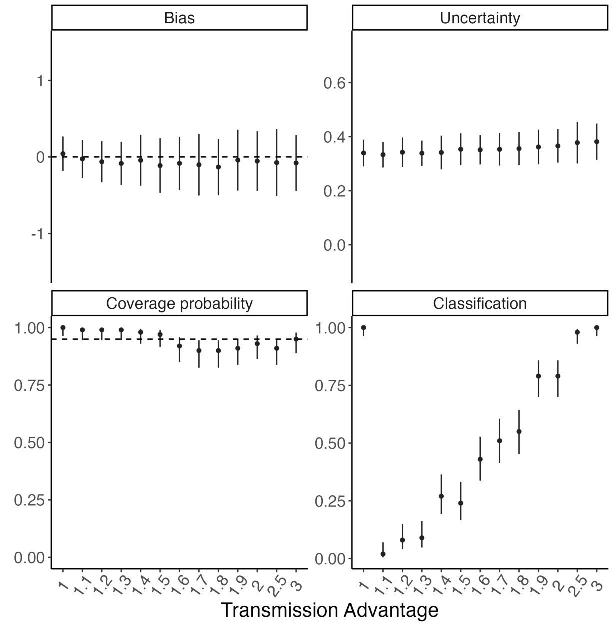

(B) $R_t$ = 1.6 and 10 days of data

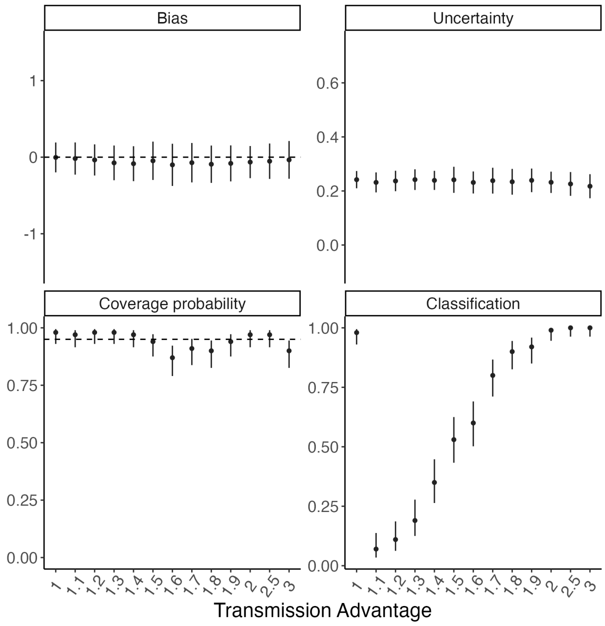

(C) $R_t = 1.1$ and 20 days of data

(D) $R_t = 1.6$ and 20 days of data

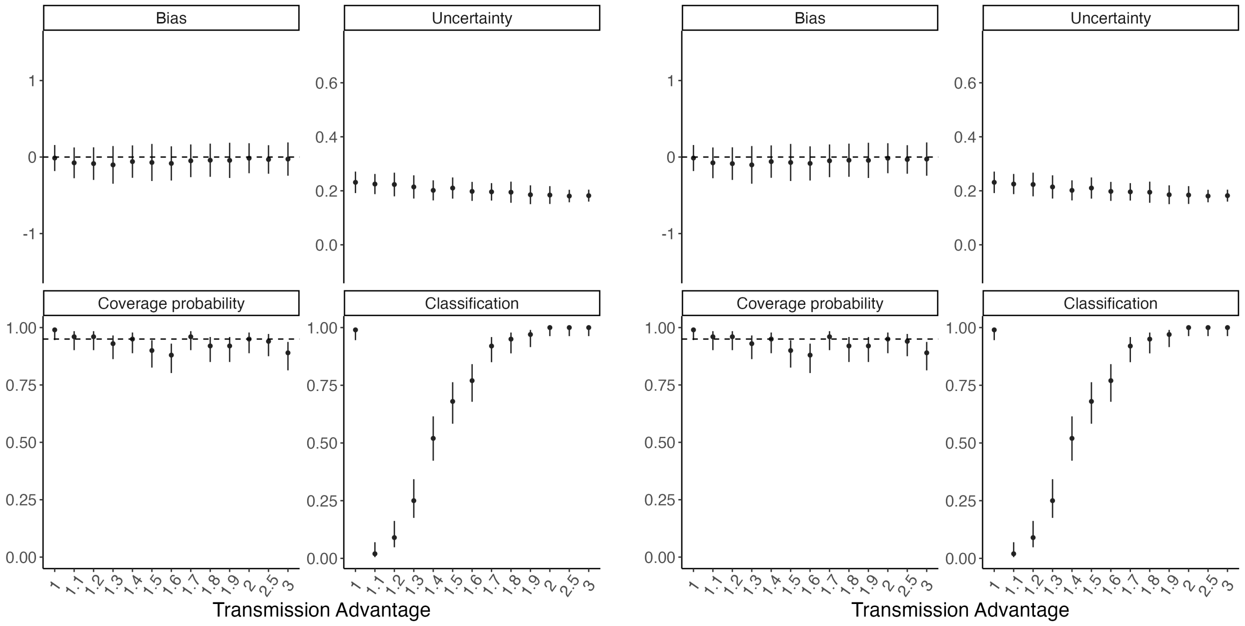

Figure S5: Method performance using simulated data assuming the same natural history for the reference and the variant (using 10 or 20 days of incidence data). In each panel, the dots and vertical bars represent the central estimate and uncertainty respectively. See Sec. 5.2 for details.

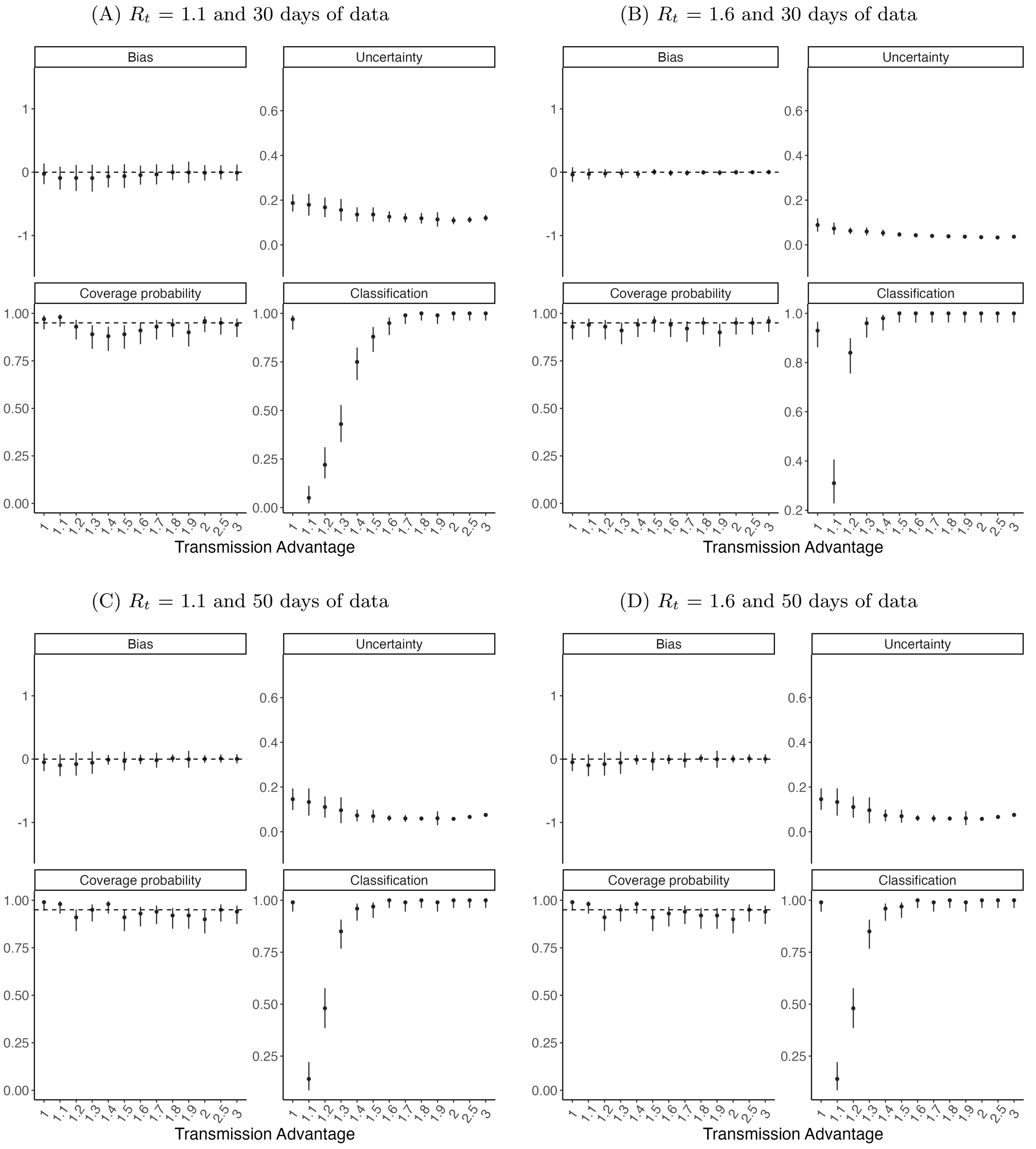

Figure S6: Method performance using simulated data assuming the same natural history for the reference and the variant (using 30 or 50 days of incidence data). In each panel, the dots and vertical bars represent the central estimate and uncertainty respectively. See Sec. 5.2 for details.

#### 5.4 Sensitivity to serial interval mean

In this section, we present results for the scenario where data were simulated assuming different natural history parameters for the reference and the variant. We assumed that the mean serial interval of the variant is $0.5, 1.5$, or 2 times that of the reference. Further, we assumed that the parameters of both the reference and the variant are correctly specified during estimations. Results are shown using $10, 20, 30$, and 50 days of incidence data.

(A) $R_t = 1.1$ and 10 days of data

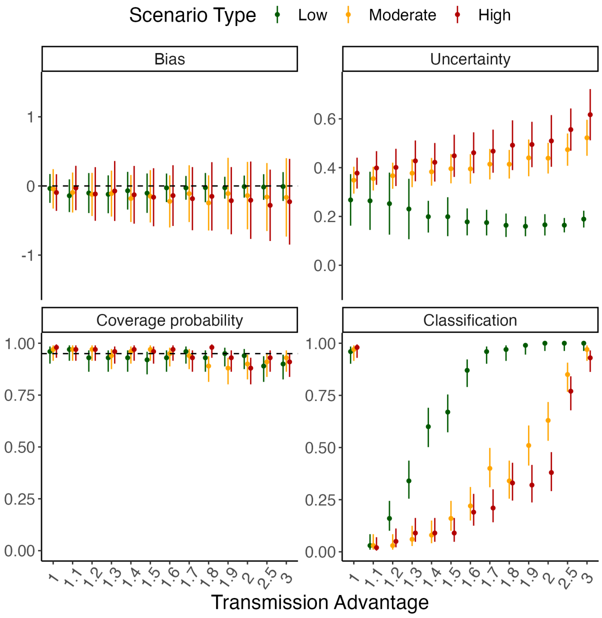

(B) $R_t = 1.6$ and 10 days of data

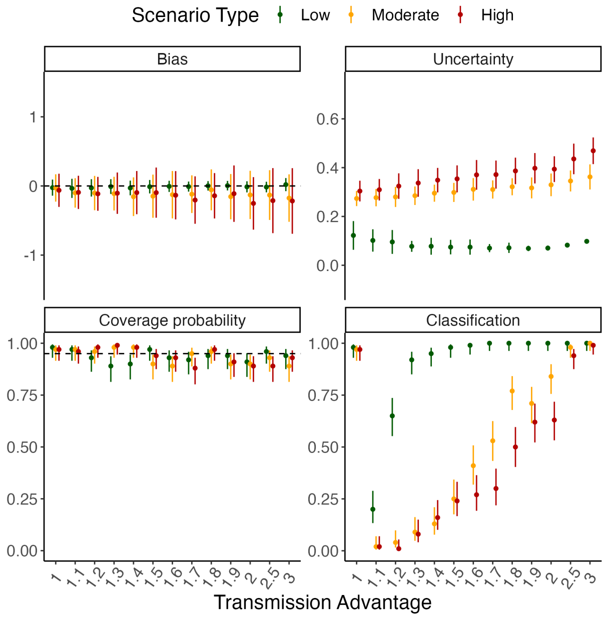

(C) $R_t$ = 1.1 and 20 days of data

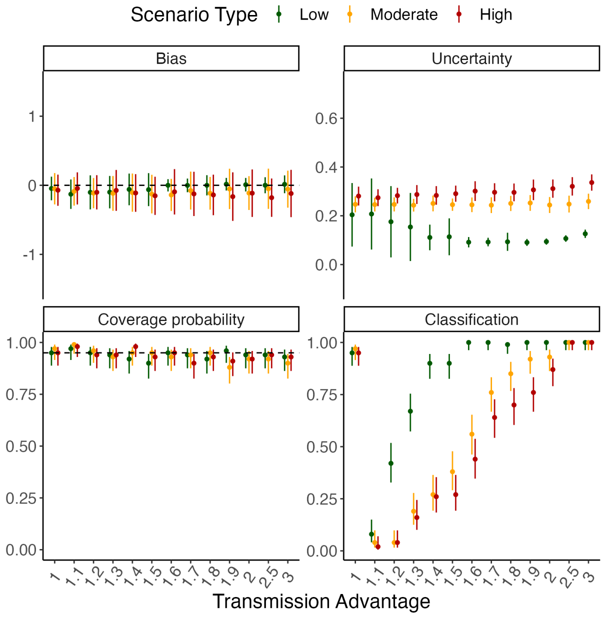

(D) $R_t$ = 1.6 and 20 days of data

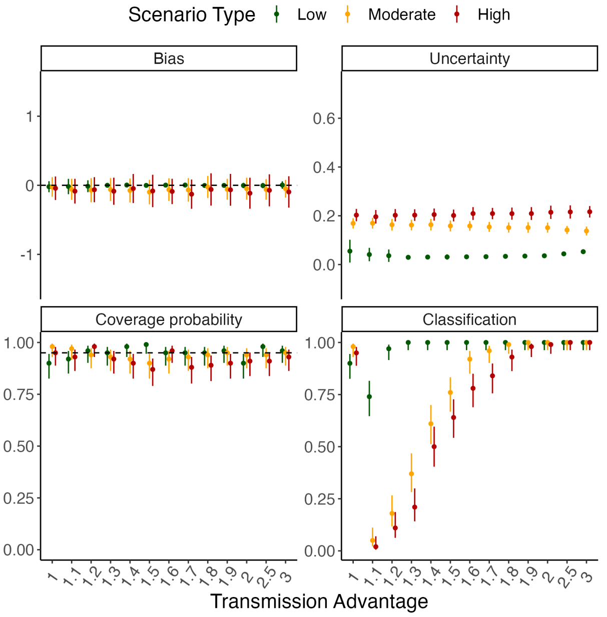

Figure S7: Method performance using simulated data assuming different SI mean for the reference and the variant (using 10 or 20 days of incidence data). The mean serial interval of the variant is assumed to be 0.5 (low), 1.5 (moderate) or 2 (high) times that of the reference. In each panel, the dots and vertical bars represent the central estimate and uncertainty respectively. See Sec. 5.2 for details.

(A) $R_t$ = 1.1 and 30 days of data

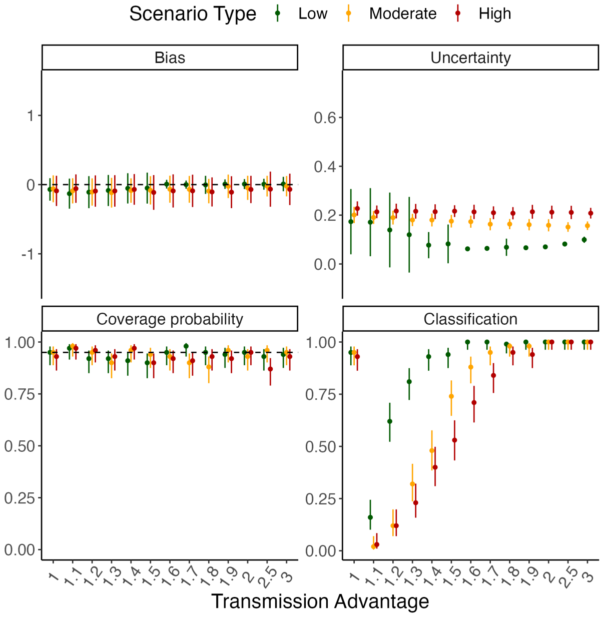

(B) $R_t$ = 1.6 and 30 days of data

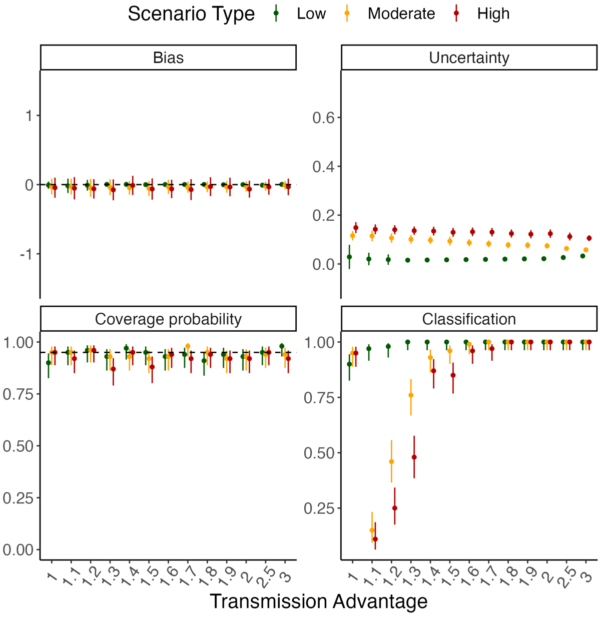

(C) $R_t$ = 1.1 and 50 days of data

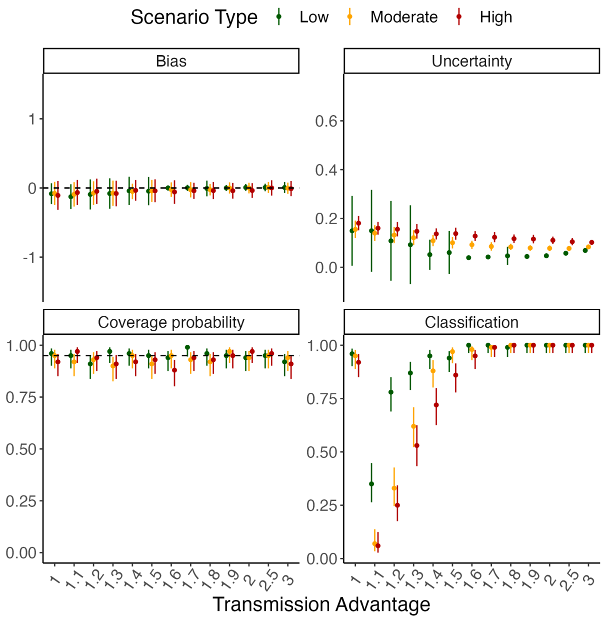

(D) $R_t$ = 1.6 and 50 days of data

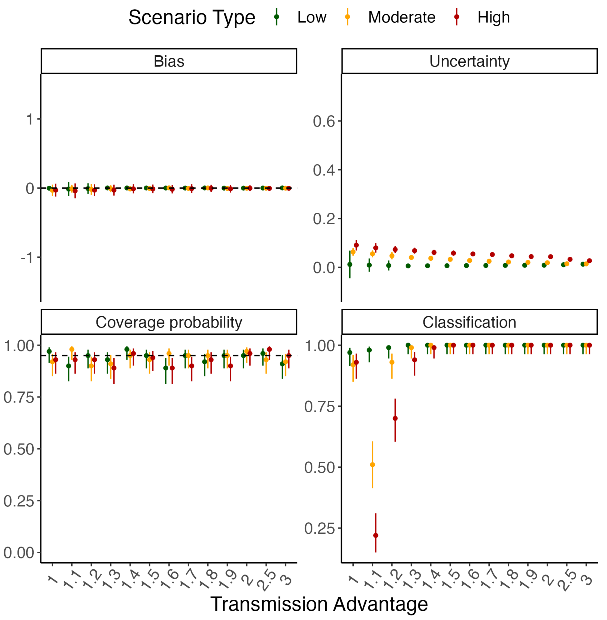

Figure S8: Method performance using simulated data assuming different SI mean for the reference and the variant (using 30 or 50 days of incidence data). The mean serial interval of the variant is assumed to be 0.5 (low), 1.5 (moderate) or 2 (high) times that of the reference. In each panel, the dots and vertical bars represent the central estimate and uncertainty respectively. See Sec. 5.2 for details.

#### 5.5 Misspecification of serial interval mean

In this section, we present results for the scenario where data were simulated assuming different natural history parameters for the reference and the variant. We assumed that the mean serial interval of the variant is 0.5, 1.5, or 2 times that of the reference. However, we assumed that the parameters of both the reference and the variant are assumed to be the same during estimation. Results are shown using 10, 20, 30, and 50 days of incidence data.

(A) $R_t = 1.1$ and 10 days of data

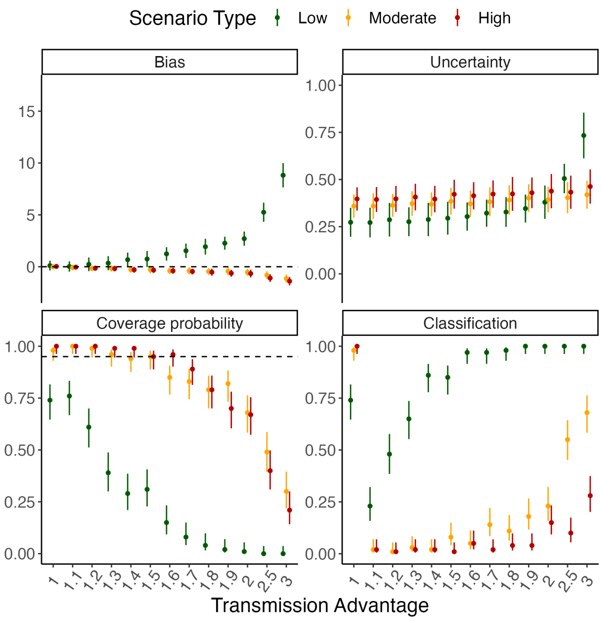

(B) $R_t = 1.6$ and 10 days of data

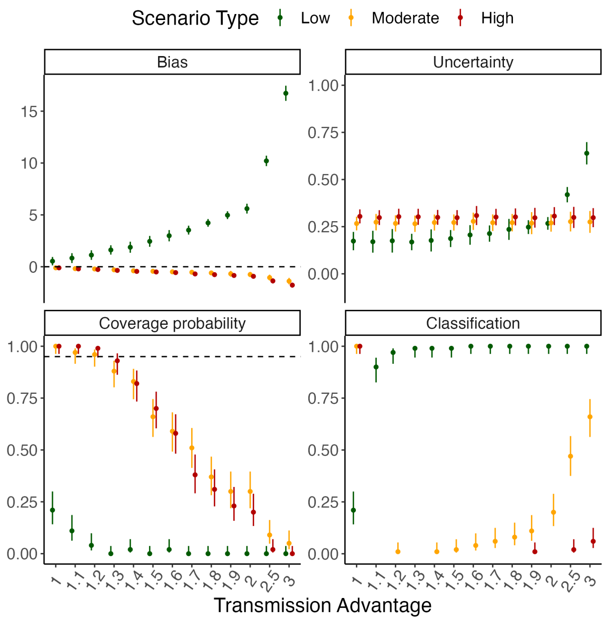

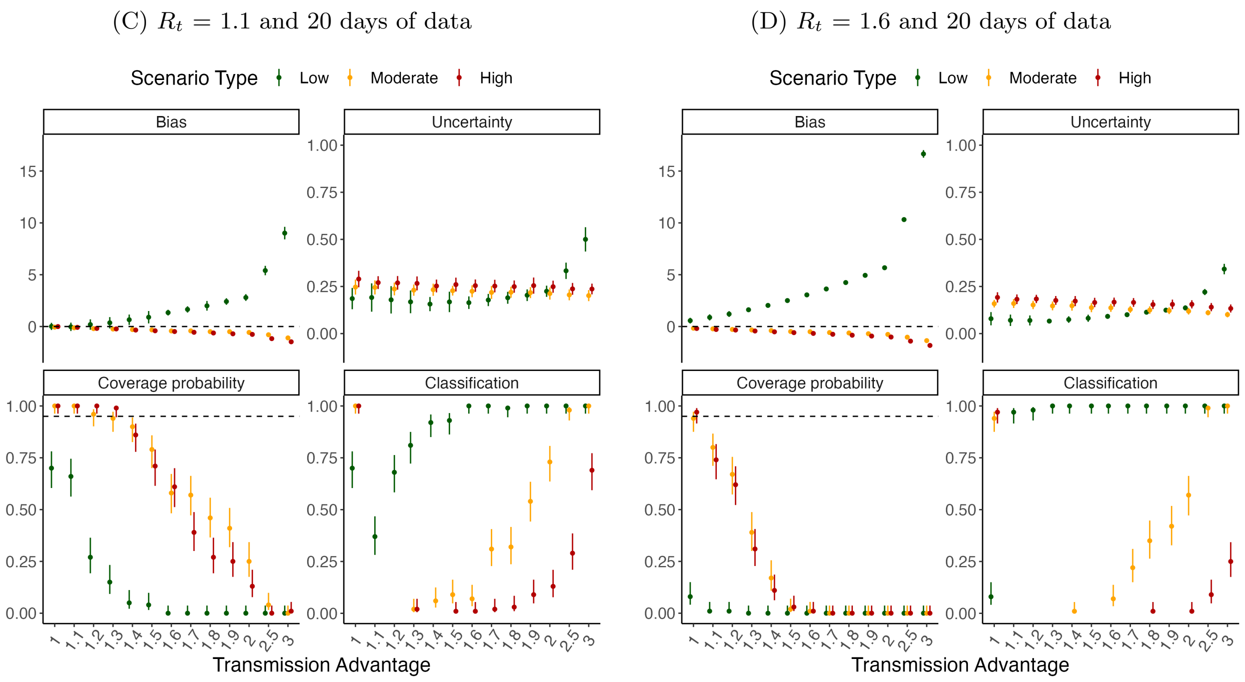

Figure S9: Method performance using simulated incidence data when the mean SI of the variant is different and is misspecified during estimation (using 10 or 20 days of incidence data). The mean serial interval of the variant is assumed to be 0.5 (low), 1.5 (moderate) or 2 (high) times that of the reference. In each panel, the dots and vertical bars represent the central estimate and uncertainty respectively. See Sec. 5.2 for details.

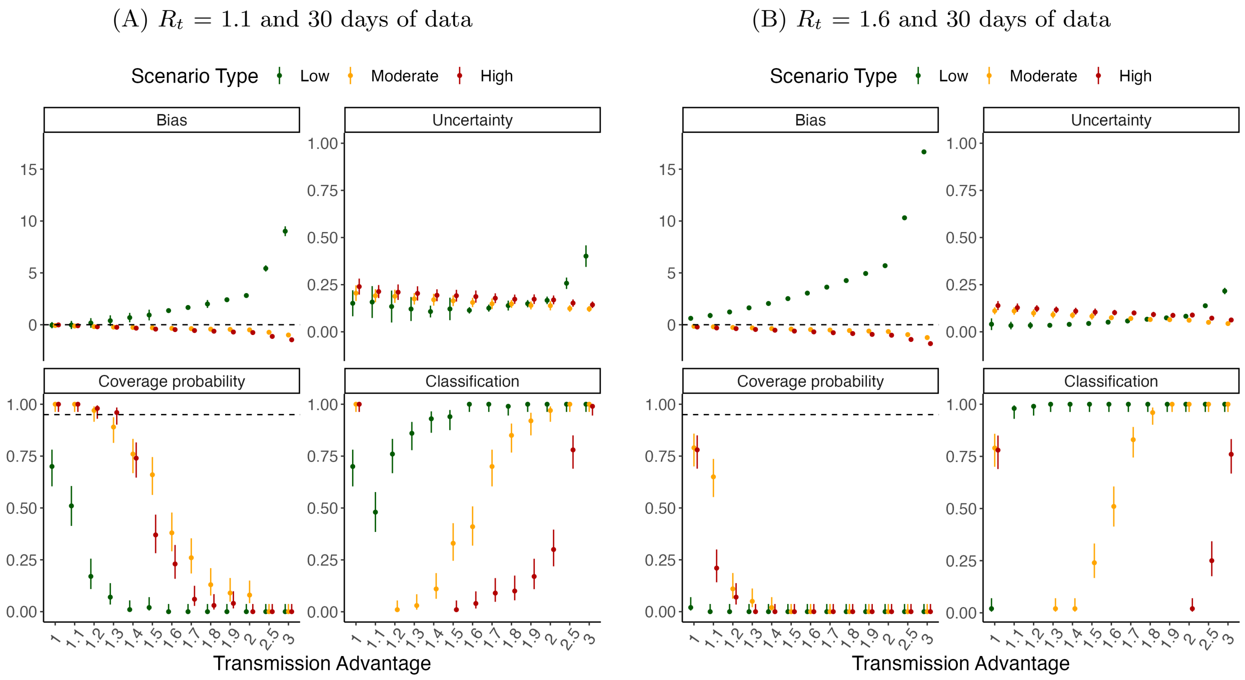

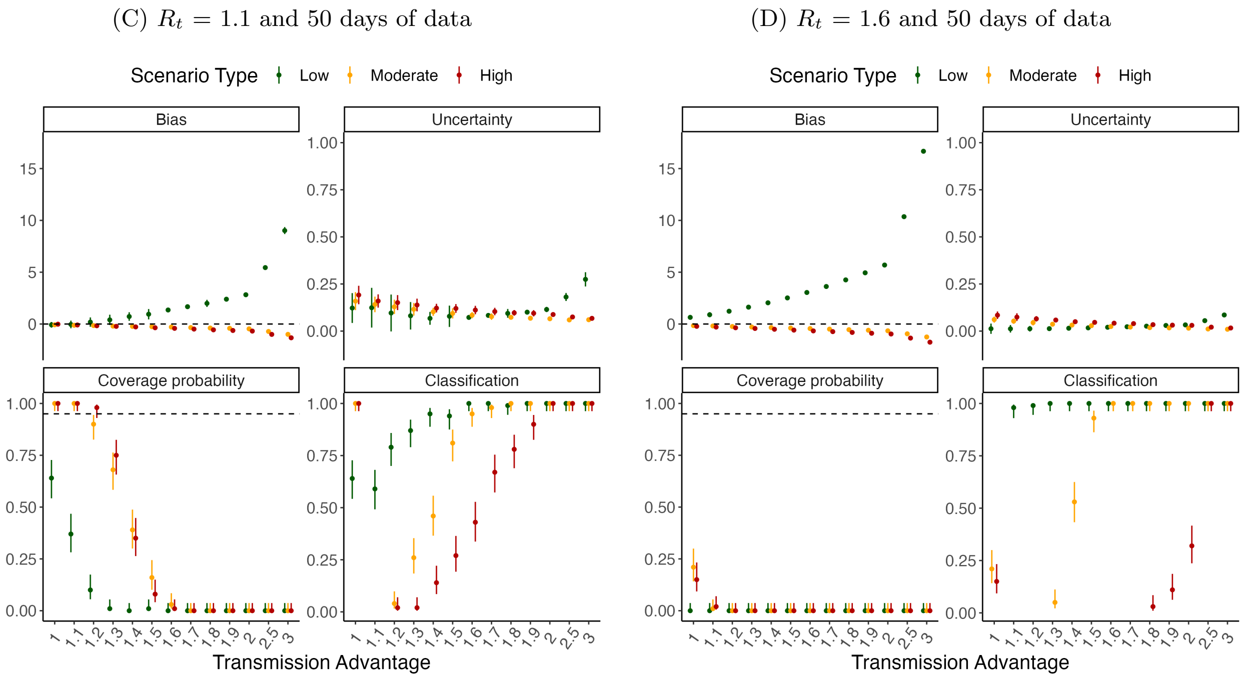

Figure S10: Method performance using simulated incidence data when the mean SI of the variant is different and is misspecified during estimation (using 30 or 50 days of incidence data). The mean serial interval of the variant is assumed to be 0.5 (low), 1.5 (moderate) or 2 (high) times that of the reference. In each panel, the dots and vertical bars represent the central estimate and uncertainty respectively. See Sec. 5.2 for details.

#### 5.6 Sensitivity to serial interval CV

In this section, we present results for the scenario where data were simulated assuming different natural history parameters for the reference and the variant. We assumed that the CV of the serial interval distribution of the variant is $0.5, 1.5$, or 2 times that of the reference. Further, we assumed that the parameters of both the reference and the variant are correctly specified during estimation. Results are shown using $10, 20, 30$, and 50 days of data.

(A) $R_t = 1.1$ and 10 days of data

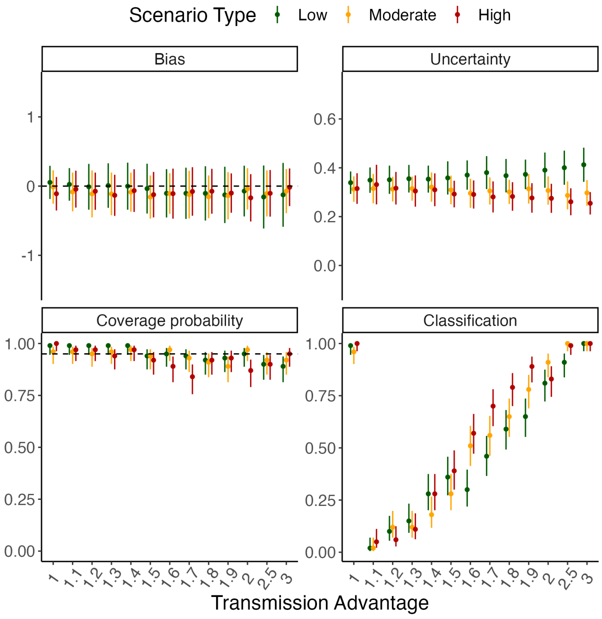

(B) $R_t = 1.6$ and 10 days of data

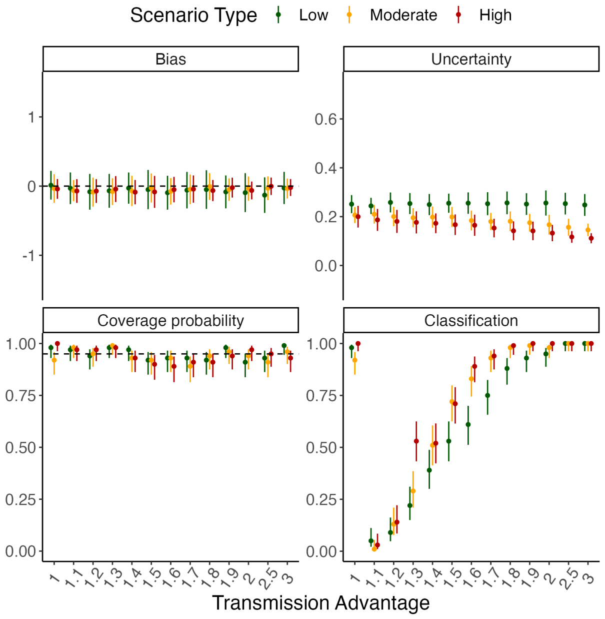

(C) $R_t$ = 1.1 and 20 days of data

(D) $R_t$ = 1.6 and 20 days of data

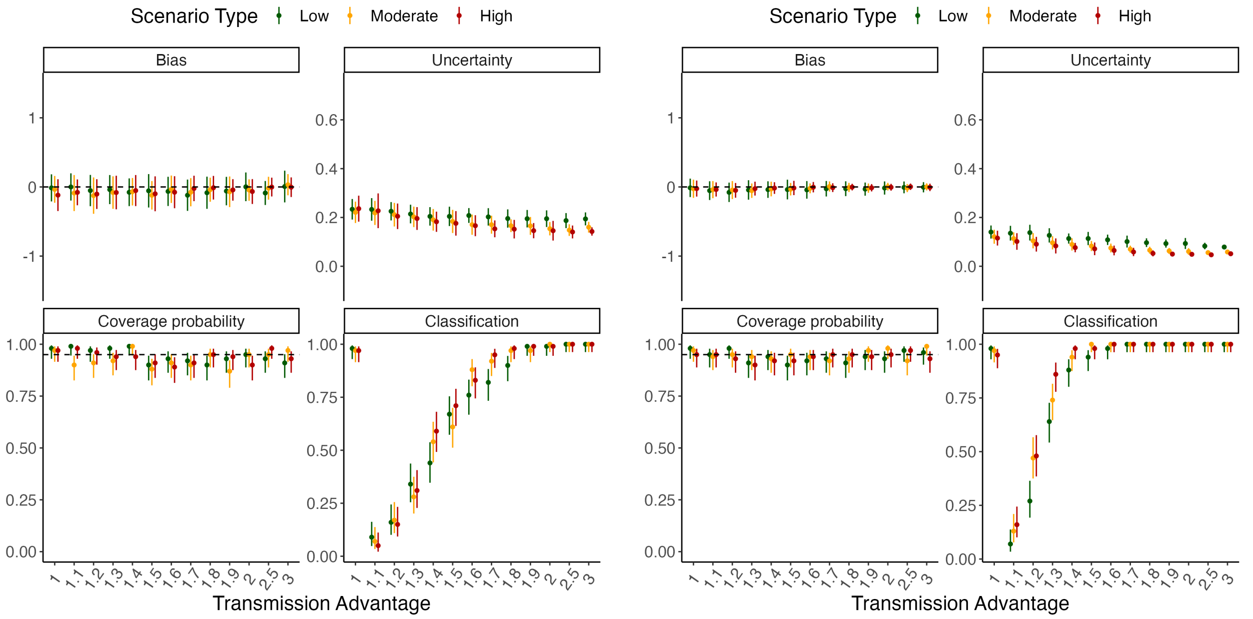

Figure S11: Method performance using simulated incidence data when the CV of the SI distribution of the variant is different and is correctly specified during estimation (using 10 or 20 days of incidence data). The CV of the serial interval distribution of the variant is assumed to be 0.5 (low), 1.5 (moderate) or 2 (high) times that of the reference. In each panel, the dots and vertical bars represent the central estimate and uncertainty respectively. See Sec. 5.2 for details.

(A) $R_t$ = 1.1 and 30 days of data

(B) $R_t$ = 1.6 and 30 days of data

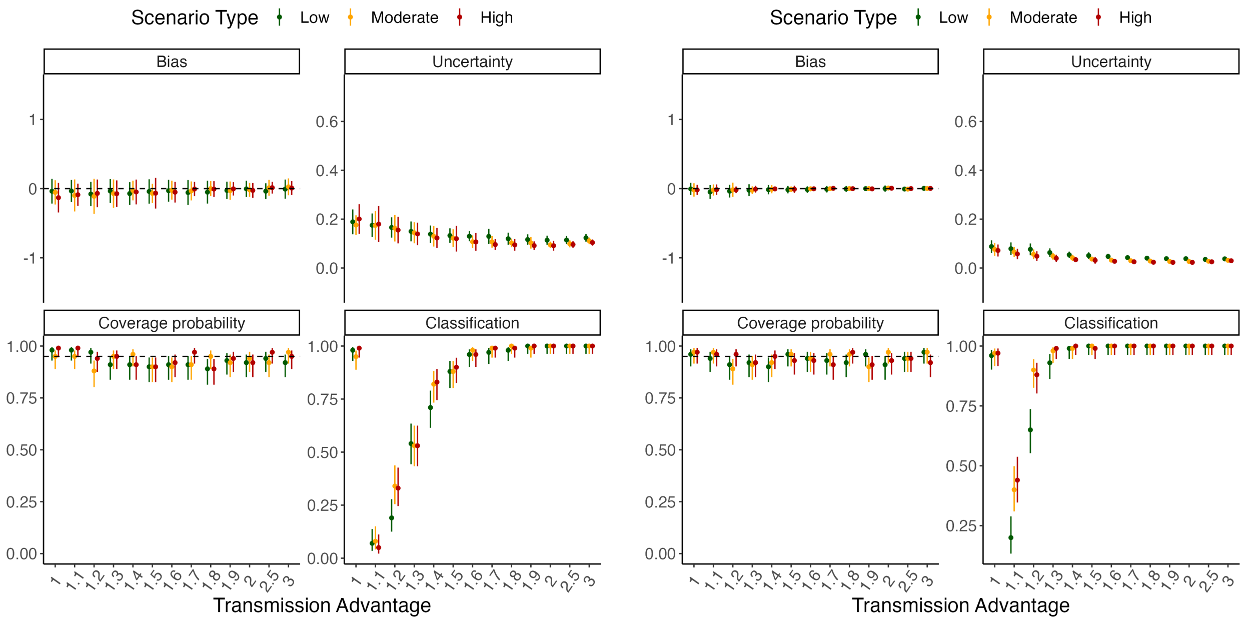

(C) $R_t = 1.1$ and 50 days of data

(D) $R_t = 1.6$ and 50 days of data

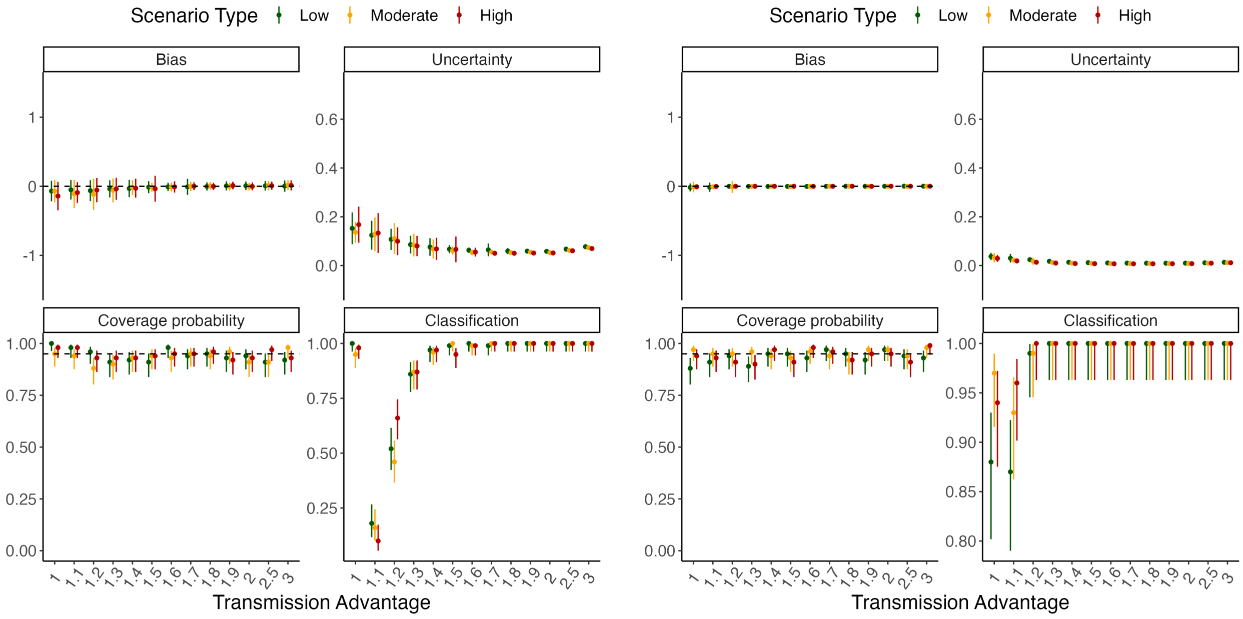

Figure S12: Method performance using simulated incidence data when the CV of the SI distribution of the variant is different and is correctly specified during estimation (using 30 or 50 days of incidence data). The CV of the serial interval distribution of the variant is assumed to be 0.5 (low), 1.5 (moderate) or 2 (high) times that of the reference. In each panel, the dots and vertical bars represent the central estimate and uncertainty respectively. See Sec. 5.2 for details.

#### 5.7 Misspecification of serial interval CV

In this section, we present results for the scenario where data were simulated assuming different natural history parameters for the reference and the variant. We assumed that the CV of the serial interval distribution of the variant is $0.5, 1.5$, or 2 times that of the reference. However, we assumed that the parameters of both the reference and the variant are assumed to be the same during estimation. Results are shown using $10, 20, 30$, and 50 days of incidence data.

(A) $R_t = 1.1$ and 10 days of data

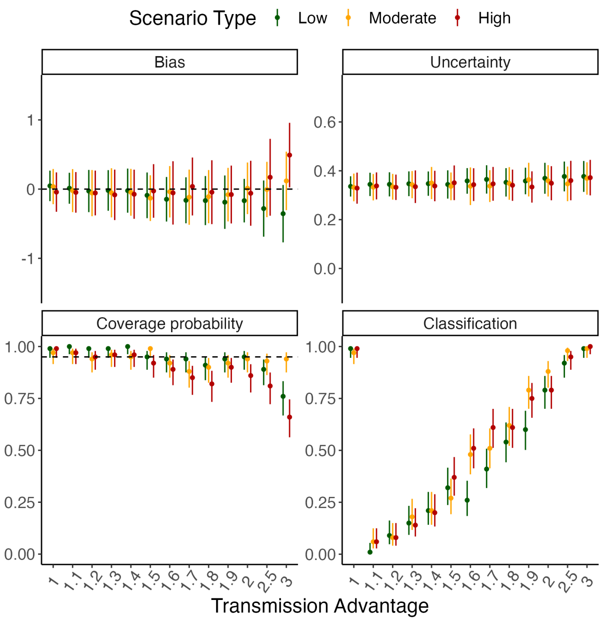

(B) $R_t = 1.6$ and 10 days of data

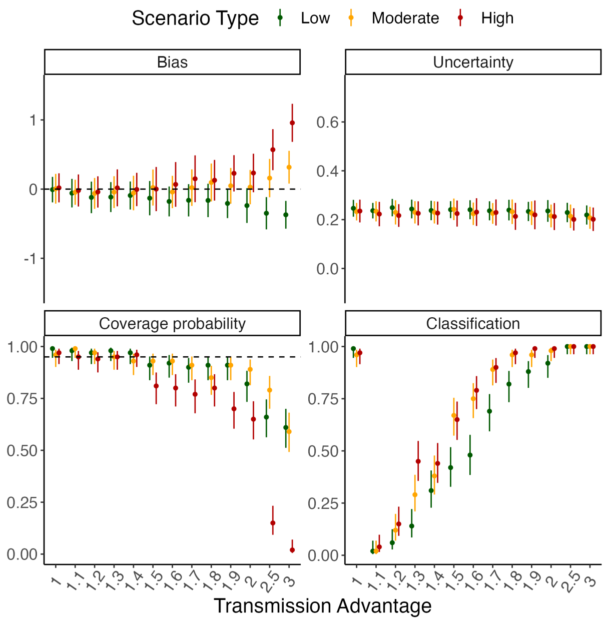

(C) $R_t = 1.1$ and 20 days of data

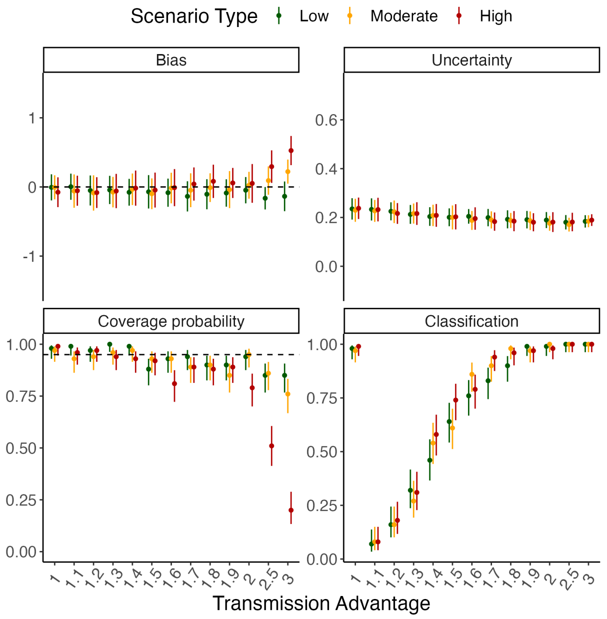

(D) $R_t = 1.6$ and 20 days of data

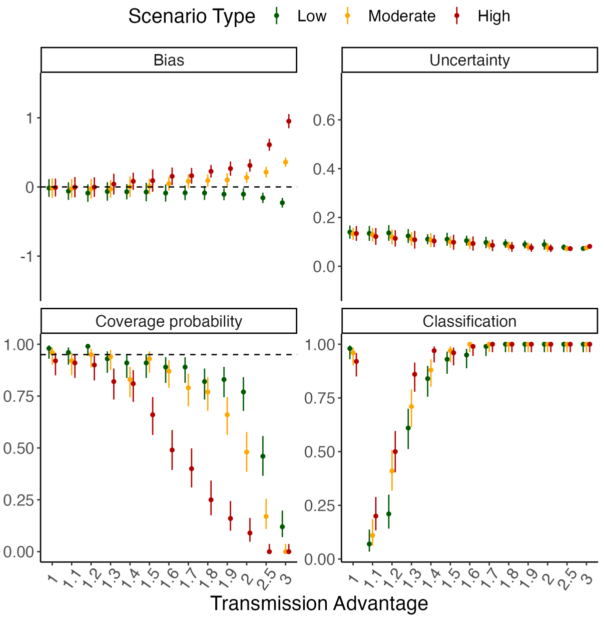

Figure S13: Method performance using simulated incidence data using simulated incidence data when the CV of the SI distribution of the variant is different and is misspecified during estimation (using 10 or 20 days of incidence data). The CV of the serial interval distribution of the variant is assumed to be 0.5 (low), 1.5 (moderate) or 2 (high) times that of the reference. In each panel, the dots and vertical bars represent the central estimate and uncertainty respectively. See Sec. 5.2 for details.

(A) $R_t = 1.1$ and 30 days of data

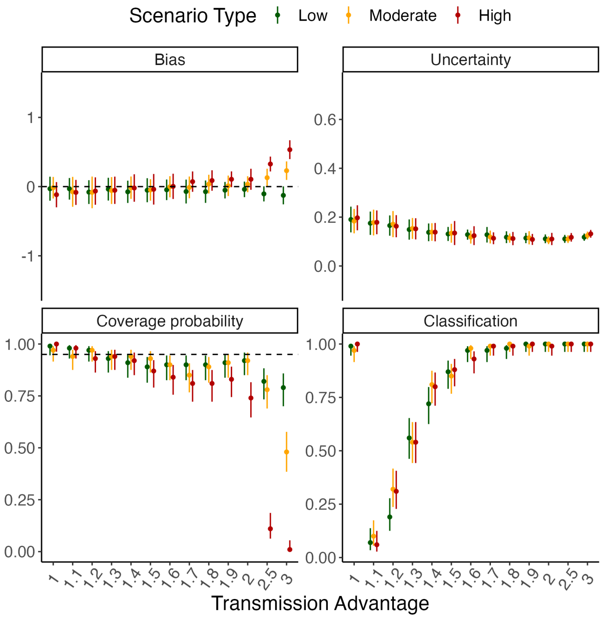

(B) $R_t = 1.6$ and 30 days of data

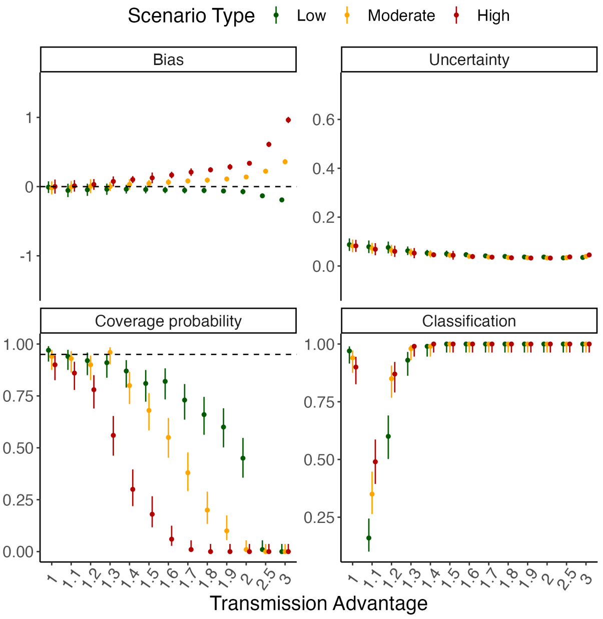

(C) $R_t$ = 1.1 and 50 days of data

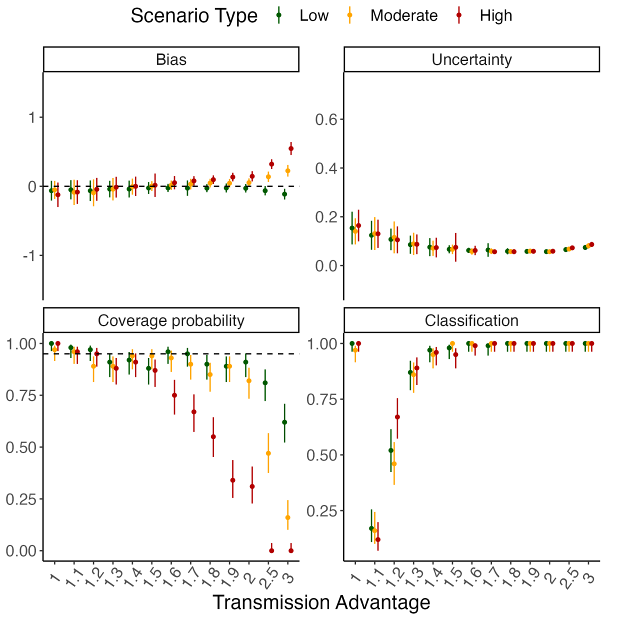

(D) $R_t$ = 1.6 and 50 days of data

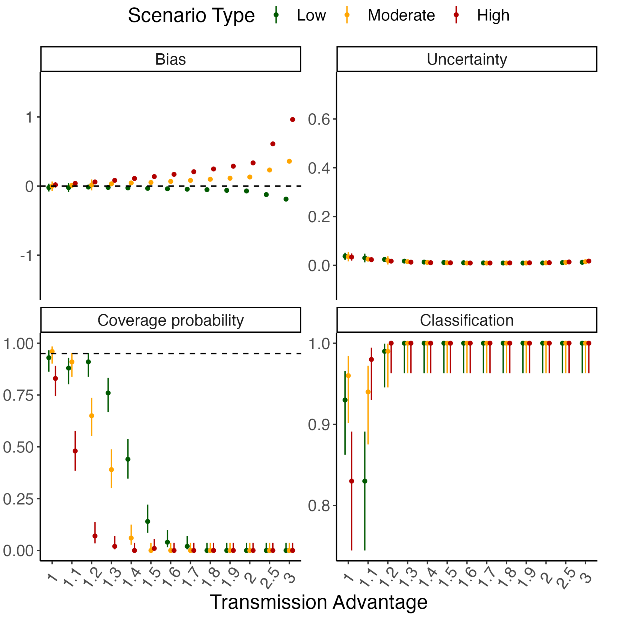

Figure S14: Method performance using simulated incidence data using simulated incidence data when the CV of the SI distribution of the variant is different and is misspecified during estimation (using 30 or 50 days of incidence data). The CV of the serial interval distribution of the variant is assumed to be 0.5 (low), 1.5 (moderate) or 2 (high) times that of the reference. In each panel, the dots and vertical bars represent the central estimate and uncertainty respectively. See Sec. 5.2 for details.

#### 5.8 Sensitivity to superspreading

To explore sensitivity of MV-EpiEstim to superspreading (which is not explicitly accounted for in MV-EpiEstim), we used a Negative Binomial distribution as the offspring distribution as in eq. (2). We simulated data using low (overdispersion parameter $\kappa = 1$), moderate ($\kappa = 0.5$), and high ($\kappa = 0.1$) levels of superspreading. This section presents the performance metrics when 10, 20, 30, and 50 days of incidence data were used for estimation.

(A) $R_t = 1.1$ and 10 days of data

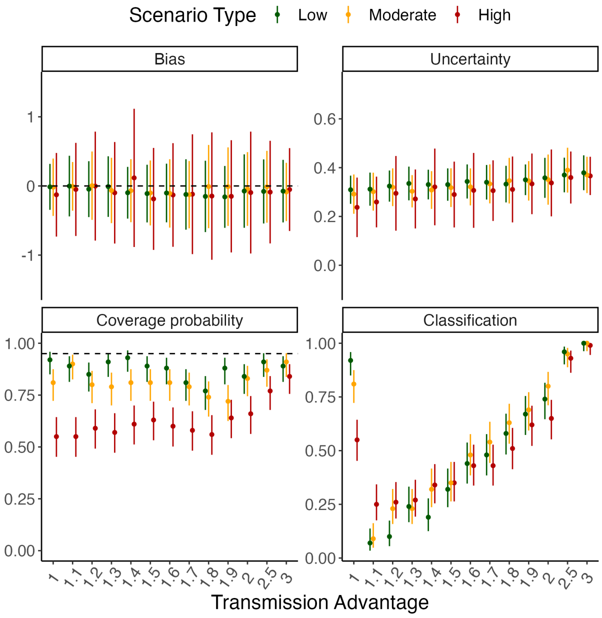

(B) $R_t = 1.6$ and 10 days of data

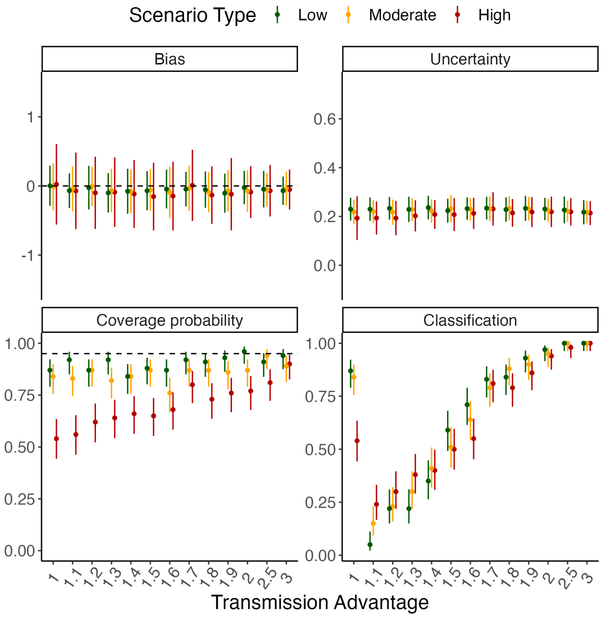

(C) $R_t$ = 1.1 and 20 days of data

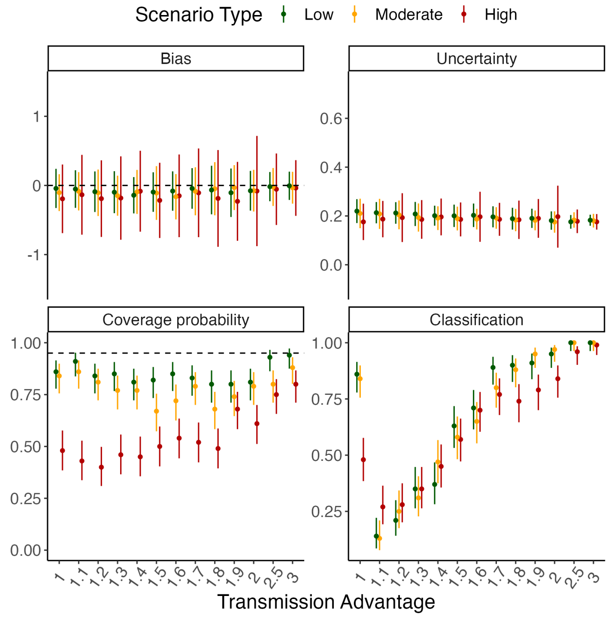

(D) $R_t$ = 1.6 and 20 days of data

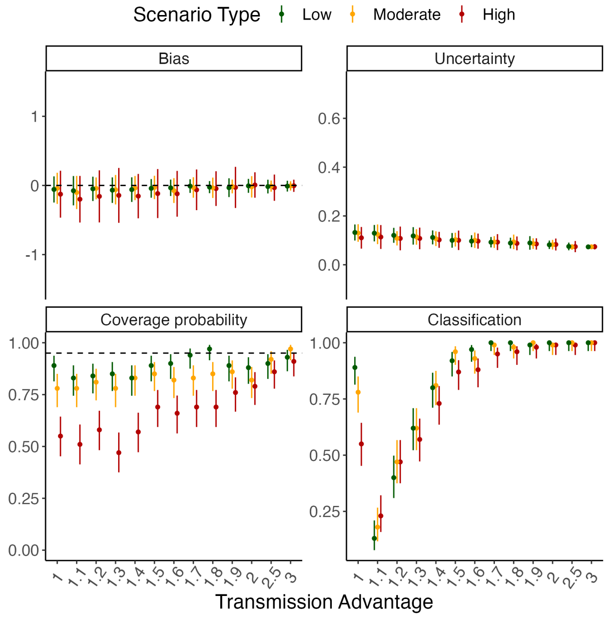

Figure S15: Method performance using incidence data simulated with superspreading (using 10 or 20 days of incidence data). We simulated data with low (overdispersion parameter $\kappa = 1$), moderate ($\kappa = 0.5$) and high ($\kappa = 0.1$) levels of superspreading. In each panel, the dots and vertical bars represent the central estimate and uncertainty respectively. See Sec. 5.2 for details.

(A) $R_t$ = 1.1 and 30 days of data

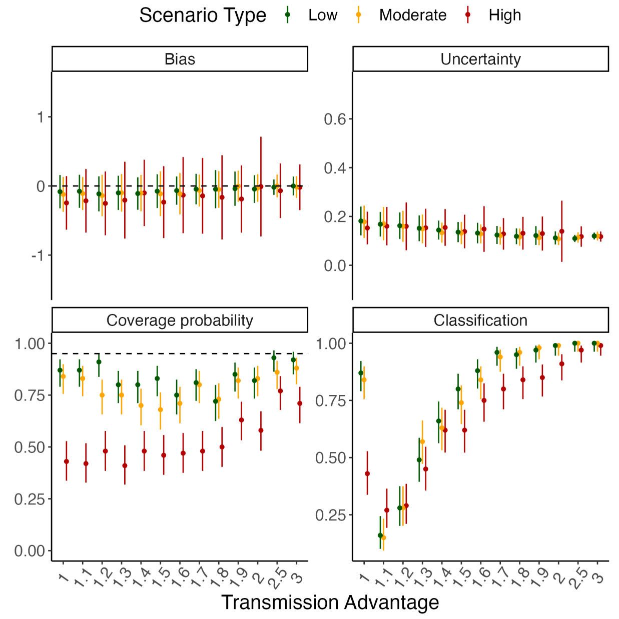

(B) $R_t$ = 1.6 and 30 days of data

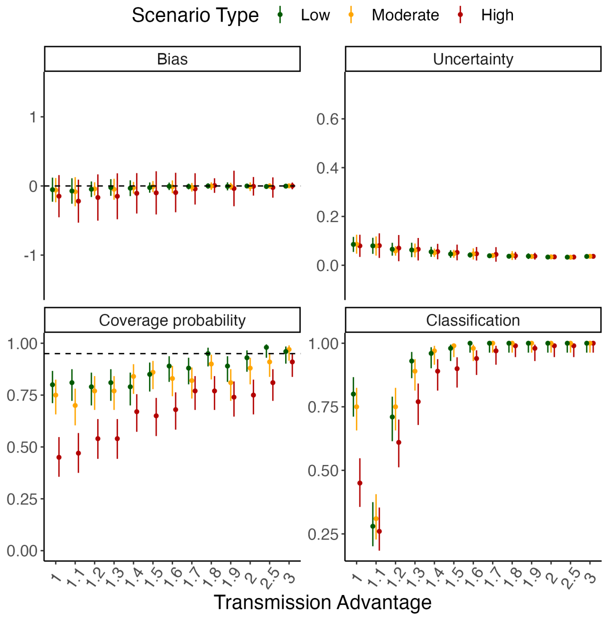

(C) $R_t = 1.1$ and 50 days of data

(D) $R_t = 1.6$ and 50 days of data

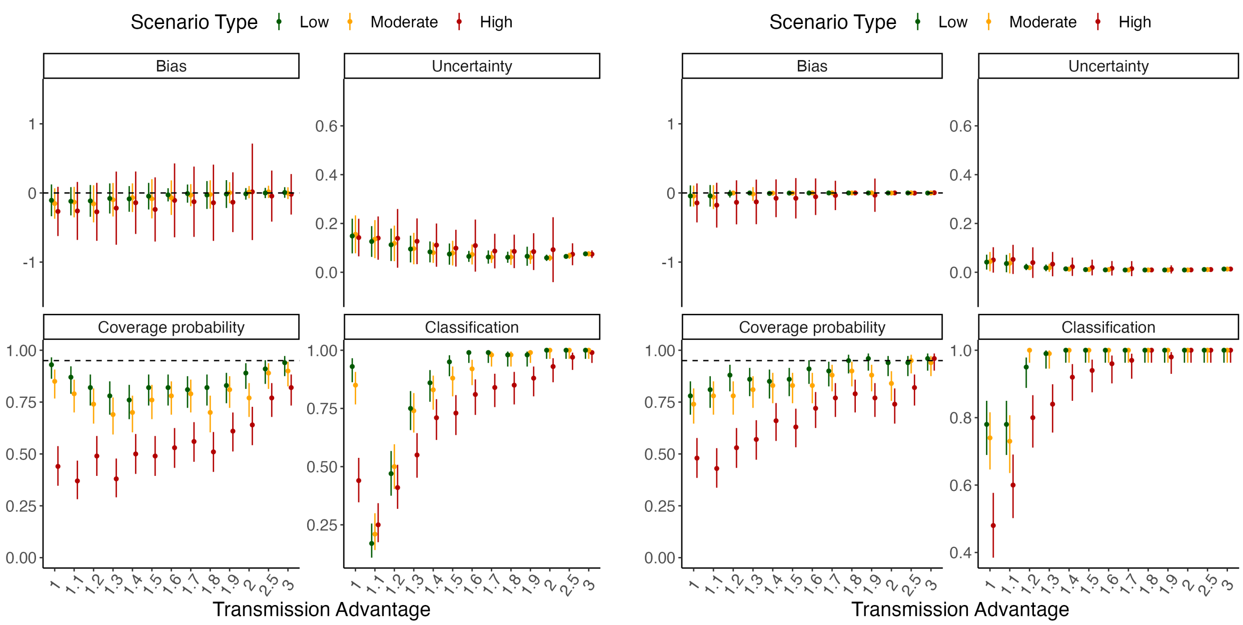

Figure S16: Method performance using incidence data simulated with superspreading (using 30 or 50 days of incidence data). We simulated data with low (overdispersion parameter $\kappa = 1$), moderate ($\kappa = 0.5$) and high ($\kappa = 0.1$) levels of superspreading. In each panel, the dots and vertical bars represent the central estimate and uncertainty respectively. See Sec. 5.2 for details.

#### 5.9 Sensitivity to under-reporting

To explore the sensitivity of our method to under-reporting, we first simulated data as in the baseline scenario (Sec. 5.3). We then assumed a constant reporting probability for both the reference and the variant and estimated the effective transmission advantage using only the reported cases. We set the reporting probability to 0.2, 0.5, or 0.8. This section presents the performance metrics when 10, 20, 30, and 50 days of incidence data were used for estimation.

(A) $R_t$ = 1.1 and 10 days of data

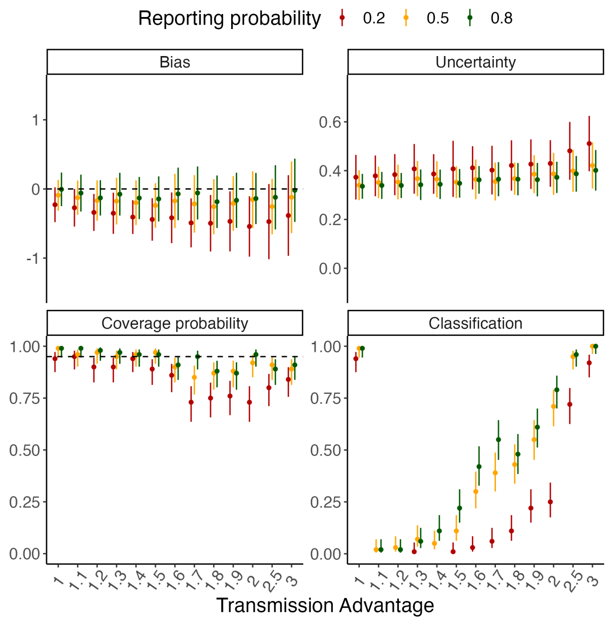

(B) $R_t$ = 1.6 and 10 days of data

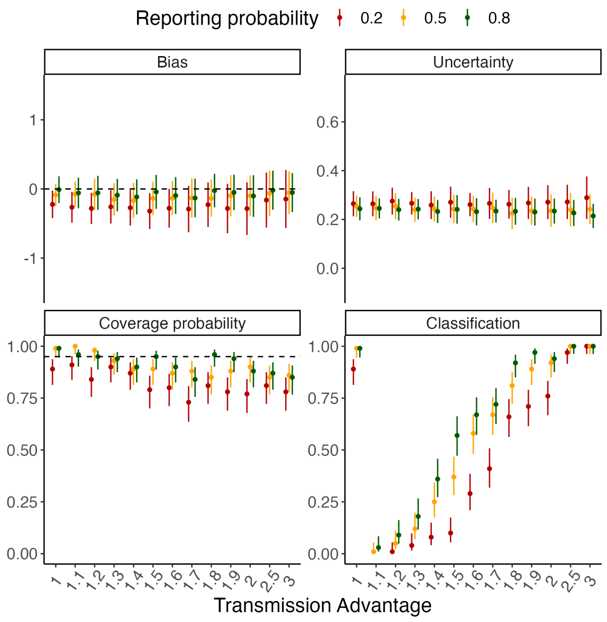

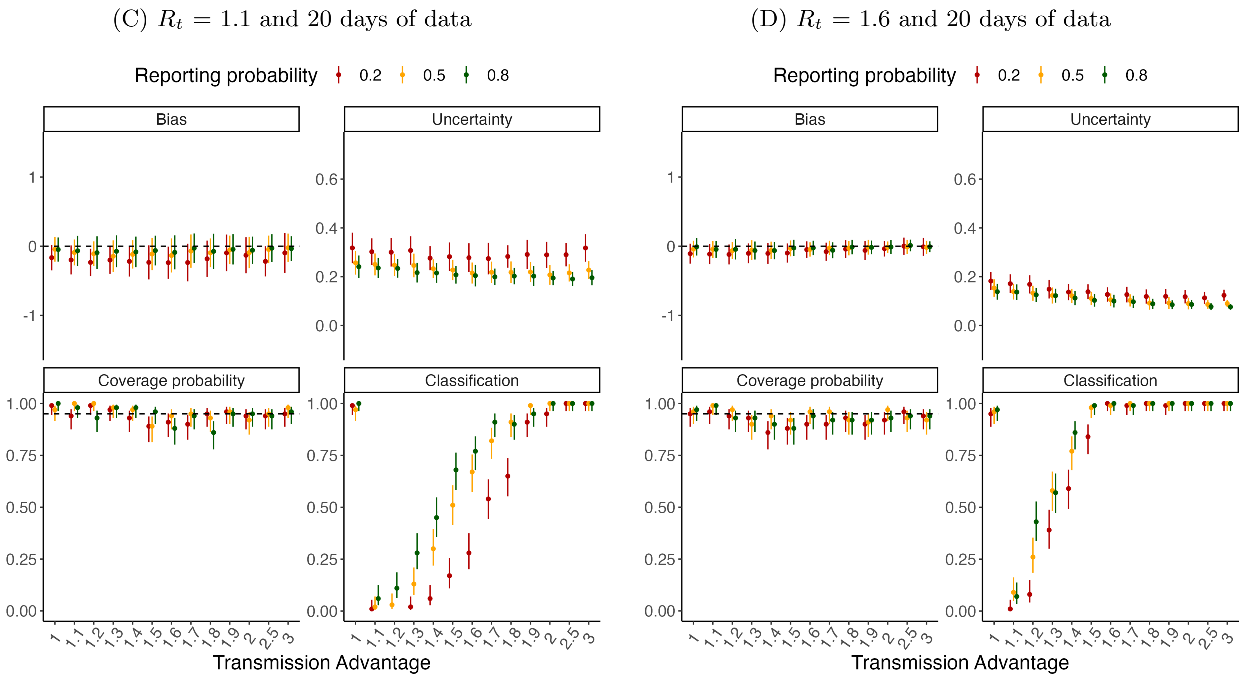

Figure S17: Method performance using using simulated incidence data with under-reporting (using 10 or 20 days of incidence data). We assume that the reporting probability is 0.2, 0.5, or 0.8. In each panel, the dots and vertical bars represent the central estimate and uncertainty respectively. See Sec. 5.2 for details.

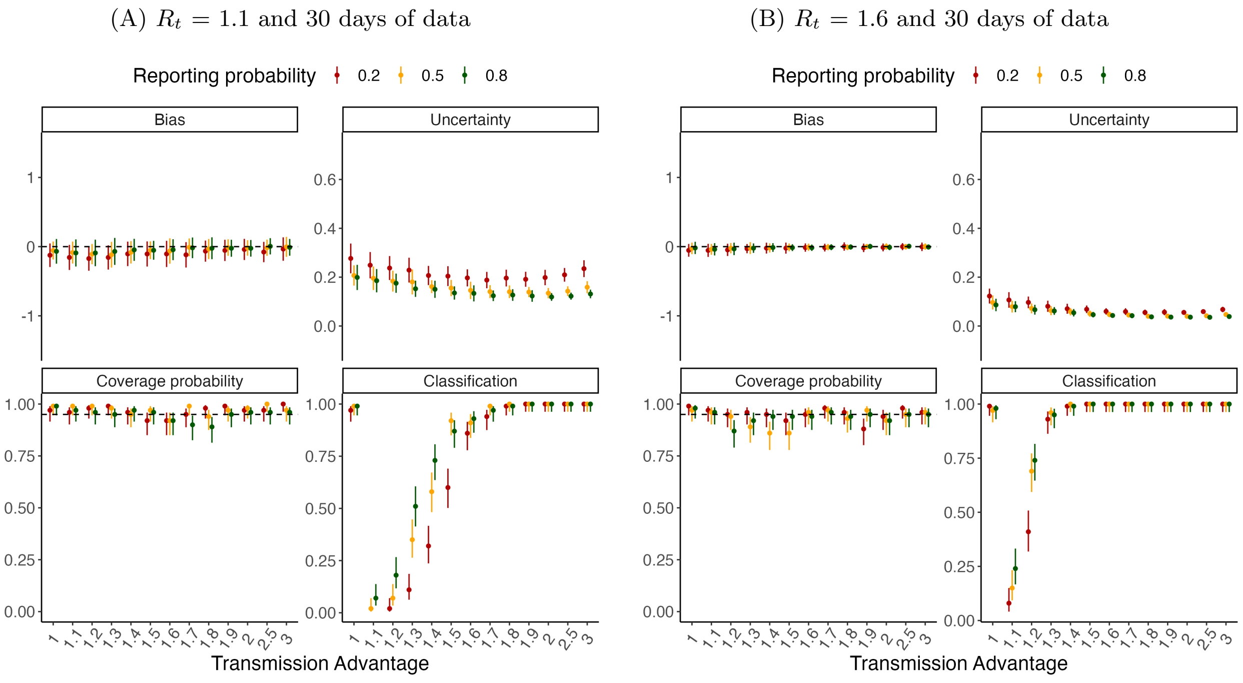

(C) $R_t$ = 1.1 and 50 days of data (D) $R_t$ = 1.6 and 50 days of data

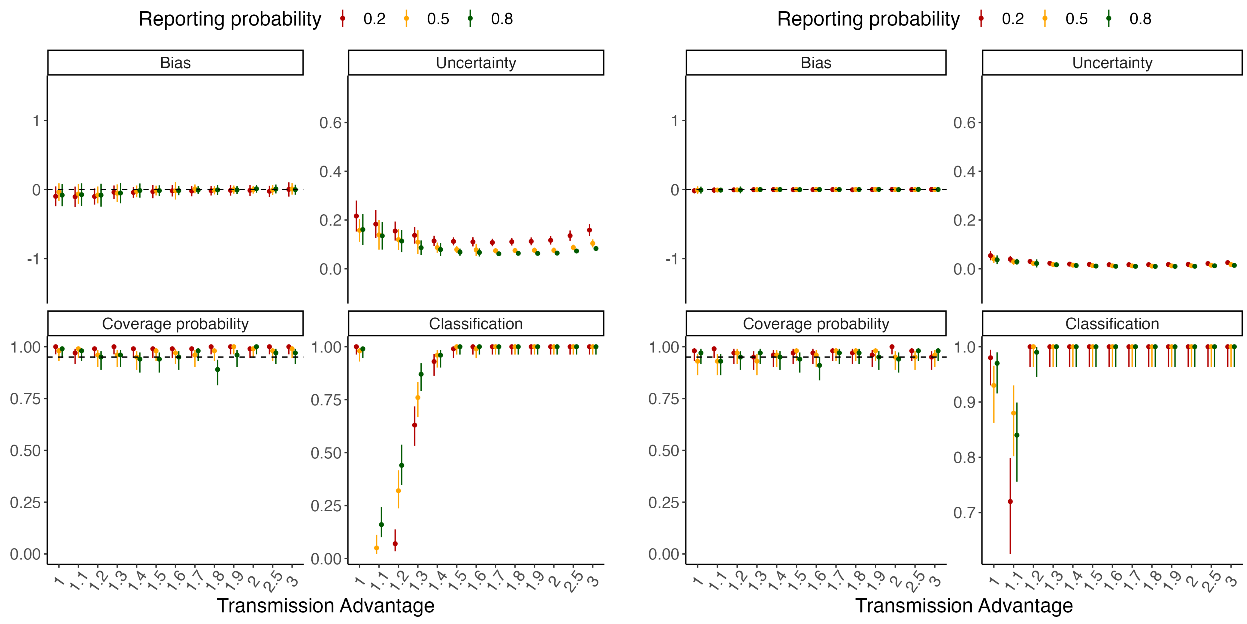

Figure S18: Method performance using using simulated incidence data with under-reporting (using 30 or 50 days of incidence data). We assume that the reporting probability is 0.2, 0.5, or 0.8. In each panel, the dots and vertical bars represent the central estimate and uncertainty respectively. See Sec. 5.2 for details.

#### 5.10 Time-varying $R_t$

To explore the effect of changing transmission dynamics on the estimates, we simulated data as in the baseline scenario (Sec. 5.3) but with the reference $R_t$ changing after 30 days from either 1.4 to 1.1, or 1.6 to 1.2. This section presents the performance metrics when 50 days of incidence data were used for estimation (i.e. covering the period both before and after the step-change in $R_t$).

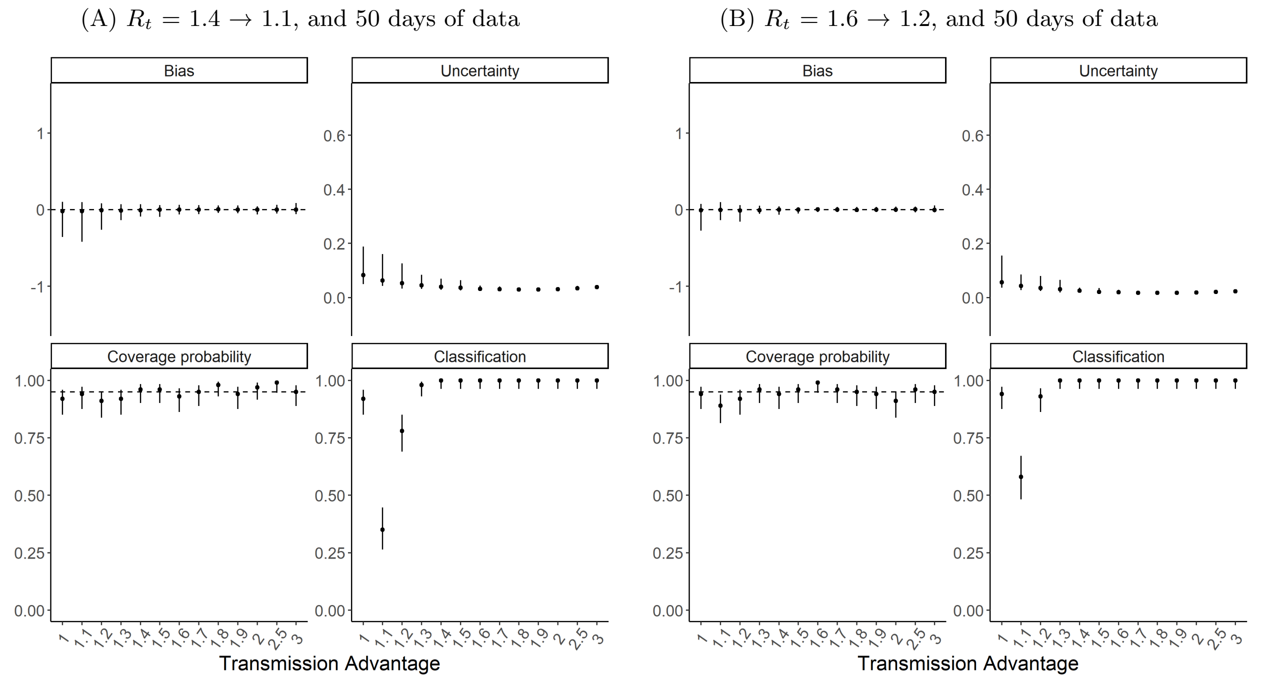

Figure S19: Method performance using incidence data simulated with time-varying $R_t$. The reference $R_t$ changes after 30 days of the simulation. Here we use 50 days of incidence data for estimation. In each panel, the dots and vertical bars represent the central estimate and uncertainty respectively. See Sec. 5.2 for details.

#### 5.11 Two locations with time-varying $R_t$

We explored the performance of our method in the presence of changing transmission dynamics when data from two locations are used for estimation, instead of a single location. We simulated independent epidemics in two locations as in the baseline scenario (Sec. 5.3) but with the $R_t$ profile changing over time. The reference $R_t$ decreased from (i) 1.4 to 1.1, or (ii) 1.6 to 1.2, after 20 days in the first location and after 40 days in the second location. We also explored a further scenario where the decrease in reference $R_t$ is different in the two locations and occurs at different times (from 1.4 to 1.1 after 40 days in the first location, and from 1.6 to 1.2 after 20 days in the second location). Tab S5 presents a summary of the $R_t$ profiles used for simulations. This section presents the performance metrics when 50 days of incidence data were used for estimation (i.e. covering the period both before and after the step-changes in $R_t$).

| Location 1 | | | Location 2 | | |
|---|---|---|---|---|---|
| Initial $R_t$ | Final $R_t$ | Time of $R_t$ change | Initial $R_t$ | Final $R_t$ | Time of $R_t$ change |
| 1.4 | 1.1 | 20 days | 1.4 | 1.1 | 40 days |
| 1.6 | 1.2 | 20 days | 1.6 | 1.2 | 40 days |
| 1.4 | 1.1 | 40 days | 1.6 | 1.2 | 20 days |

Table S5: Reference $R_t$ values used to simulate incidence data in scenarios with time-varying $R_t$ profiles. The method performance results using incidence data generated by the $R_t$ profiles in rows 1, 2 and 3 are shown in fig. S20A, fig. S20B and Fig S21 respectively.

(A) $R_t = 1.4 \rightarrow 1.1$, and 50 days of data

(B) $R_t = 1.6 \rightarrow 1.2$, and 50 days of data

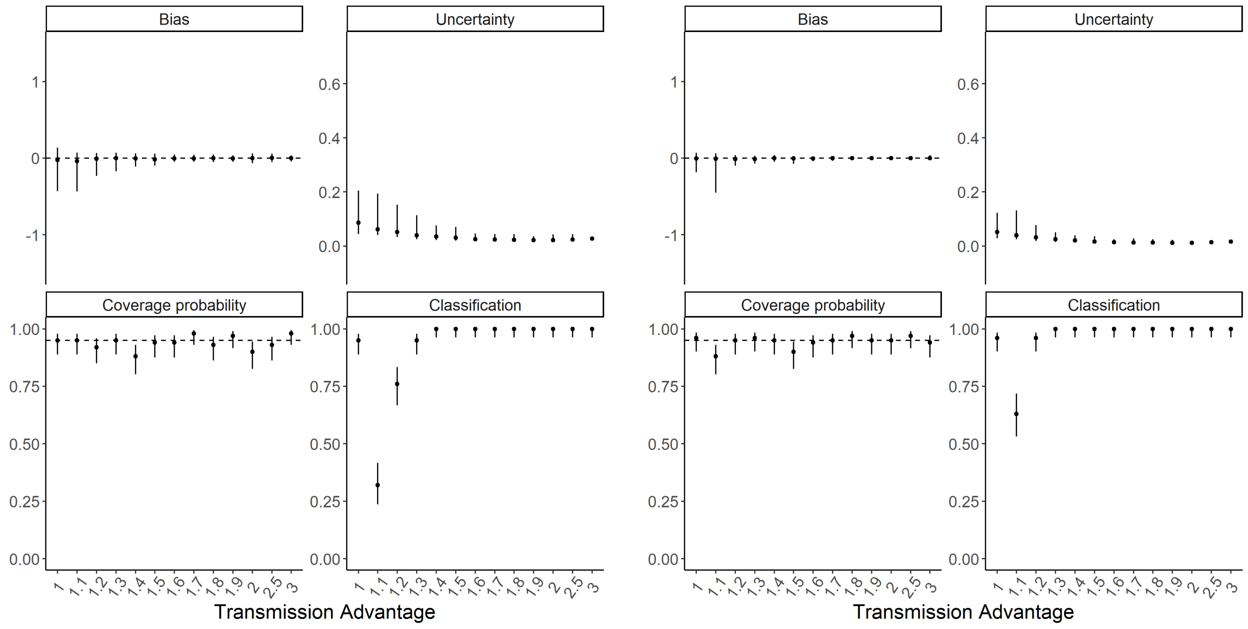

Figure S20: Method performance using incidence data simulated with time-varying $R_t$ in two locations. In both locations the reference $R_t$ decreases in the same way, but the change occurs at day 20 of the simulation for the first location location and day 40 for the second location. Here we use 50 days of incidence data for estimation. In each panel, the dots and vertical bars represent the central estimate and uncertainty respectively. See Sec. 5.2 for details.

(A) Location 1: $R_t = 1.4 \rightarrow 1.1$, Location 2: $R_t = 1.6 \rightarrow 1.2$, and 50 days of data

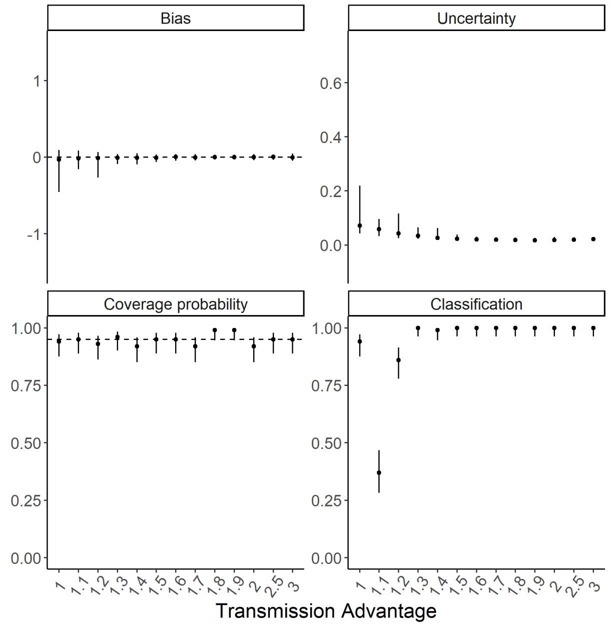

Figure S21: Method performance using incidence data simulated with time-varying $R_t$ in two locations. The reference $R_t$ decreases at day 20 in the first location and day 40 in the second location. Here we use 50 days of incidence data for estimation. In each panel, the dots and vertical bars represent the central estimate and uncertainty respectively. See Sec. 5.2 for details.

#### 6 Code and Data availability

All data and code used in this analysis are available at `https://github.com/mrc-ide/epiestims`. MV-EpiEstim is available in the development version of EpiEstim at `https://github.com/mrc-ide/EpiEstim`.
